# Risk of Post-COVID Conditions among adolescents and adults who received nirmatrelvir-ritonavir for acute COVID-19: a retrospective cohort study

**DOI:** 10.1101/2025.05.30.25327809

**Authors:** Alexandra F. Dalton, Sarah Baca, Julia Raykin, Cria O. Gregory, Tegan Boehmer, Emilia H. Koumans, Priti R. Patel, Pragna Patel, Sharon Saydah

## Abstract

**Introduction:** Post-COVID Conditions (PCC) potentially affect millions of people, but it is unclear whether treating acute COVID-19 with nirmatrelvir-ritonavir may reduce the risk of PCC.

**Methods:** Retrospective cohort study using real-world, closed claims data to assess the relationship between nirmatrelvir-ritonavir and PCC by age group (12-17, 18-49, 50-64, ≥65 years). Eligible patients had a COVID-19 index date (positive laboratory test, ICD-10 diagnosis code, or nirmatrelvir-ritonavir prescription) from April 1 – August 31, 2022, in the outpatient, telehealth, or emergency department setting, and had a higher risk of severe COVID-19 based on age (≥50 years) or underlying risk factors. Treated patients (i.e., received a nirmatrelvir-ritonavir prescription within +/-5 days of index date) were matched 1:2 on age, sex, month of index date, and HHS region with untreated patients. PCC was defined by the presence of ≥1 of 45 new-onset symptoms or conditions recorded ≥60 days after index date.

**Results:** 291,433 treated patients were matched to 582,866 untreated patients. Treatment with nirmatrelvir-ritonavir reduced PCC risk in adults 50-64 years (aHR 0.93, 95%CI 0.92-0.95) and ≥65 years (aHR 0.88, 95% CI 0.87-0.90). Treatment had minimal effect among high-risk adults 18-49 years (aHR 0.98, 95% CI 0.97-0.99), and no effect among high-risk adolescents 12-17 years (aHR 1.06, 95% CI 0.66-1.13).

**Conclusion:** Results using real-world data suggest a protective relationship between nirmatrelvir-ritonavir during acute illness and PCC risk among older adults, but not among adolescents. Consideration may be given to outpatient treatment of mild to moderate COVID-19 with nirmatrelvir-ritonavir to reduce the risk of severe disease and PCC.

## Introduction

Post-COVID Conditions (PCC) or Long COVID occur when the effects of SARS-CoV-2 infection linger weeks or months after acute illness and is associated with over 200 symptoms and conditions impacting multiple organ systems.^1^ Commonly reported symptoms include fatigue, post-exertional malaise, heart palpitations, shortness of breath and brain fog; commonly reported new-onset conditions include asthma, diabetes, and cardiovascular disease.^1^ It is estimated that among the U.S. general population, 6.9% of adults^2^ and 1.3% of children^3^ have ever had Long COVID.

Nirmatrelvir-ritonavir (Paxlovid™) is an antiviral used to treat acute mild-to-moderate COVID-19. It is authorized for persons aged 12-17 years and approved for adults ≥18 years at higher risk of progression to severe COVID-19, including adults ≥50 years, persons who are unvaccinated or not up-to-date on their COVID-19 vaccination, and patients with certain underlying medical conditions, especially immunosuppression.^4^ Studies have shown that nirmatrelvir-ritonavir reduces the severity of infection,^5,6^ the risk of hospitalization,^5,7-13^ and the risk of death including during Omicron predominance.^5,6,8,10-12^

However, it is unclear whether treating acute COVID-19 with nirmatrelvir-ritonavir may reduce the risk of PCC. Previous studies have shown mixed results^13-20^ and have not explored whether nirmatrelvir-ritonavir is differentially related to PCC across all authorized ages or presence of underlying conditions – both risk factors for severe COVID-19 and PCC.

This analysis used a large database of real-world U.S. electronic healthcare data to assess the relationship between nirmatrelvir-ritonavir and the occurrence of PCC by age group among adolescents aged 12-17 years and adults aged 18-49, 50-64, and ≥65 years.

## Methods

### Study design

We conducted a retrospective cohort study of adolescents and adults to compare the relationship between nirmatrelvir-ritonavir for treatment of acute COVID-19 and the occurrence of PCC. We used CDC-licensed HealthVerity (https://healthverity.com) closed payer claims data, which included approximately 107 million patients, of whom approximately 60% were commercially insured, 35% covered by Medicaid, and 5% covered by Medicare Advantage from April 1 – December 31, 2022 (July 2023 data release).

### Study population

Eligible patients were ≥12 years, with evidence of COVID-19 between April 1 – August 31, 2022, in the outpatient, telehealth, or emergency department (ED) setting, excluding ED visits that led to hospital admission (Figure 1). COVID-19 diagnosis was based on ICD-10 diagnosis code, positive laboratory test for SARS-CoV-2, or receipt of a nirmatrelvir-ritonavir prescription; a sensitivity analysis restricted evidence of COVID-19 to ICD-10 diagnosis code or positive laboratory test. Eligible persons had a higher risk of progression to severe COVID-19 based on age or underlying risk factors or medical conditions. Adults ≥50 years were considered at higher risk based on age alone; adolescents and adults <50 years were considered at higher risk if they had one or more select underlying risk factors (Supplemental Table 1).^21^

**Figure 1.**
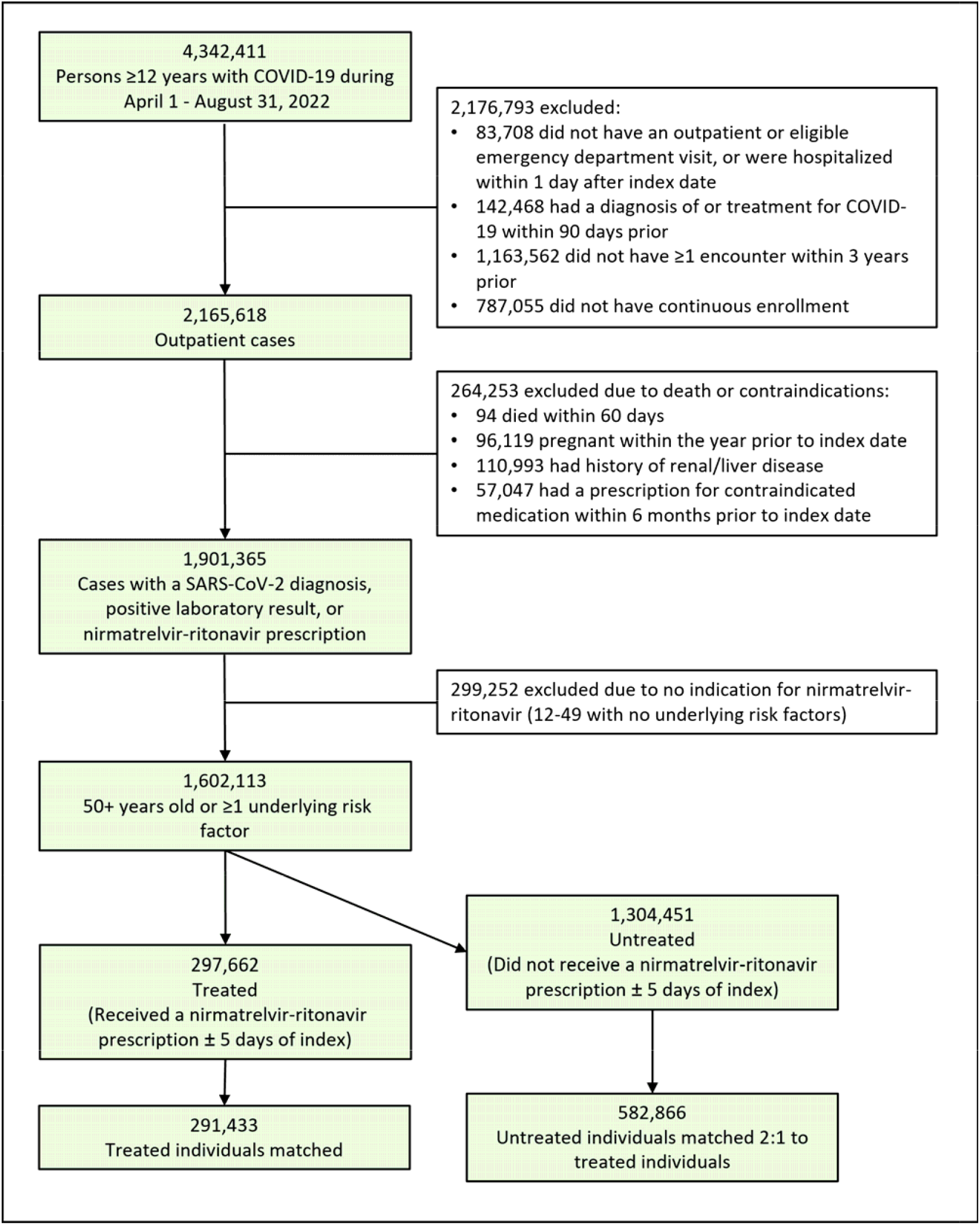
CONSORT diagram outlining inclusion and exclusion criteria to determine final analytic sample, HealthVerity, April 1 to December 31, 2022.

Patients without evidence of any healthcare encounter in the 3 years prior to their index data or lack of continuous insurance coverage 365 days before their index date and through their follow-up period were excluded. Patients who were hospitalized for COVID-19 on their index date or died within 60 days of their index date; who were pregnant within the previous year; or who were ineligible for nirmatrelvir-ritonavir due to a history of renal or liver disease, or a prescription for contraindicated medication within the previous 6 months, were excluded.

### Exposure and Outcomes

COVID-19 index date was determined by looking first for the date of a positive laboratory test for SARS CoV-2, then for an ICD-10 diagnosis code, and lastly for receipt of a nirmatrelvir-ritonavir prescription (main analysis). Treated patients were defined as having received a nirmatrelvir-ritonavir within +/-5 days of index date; untreated patients did not receive a nirmatrelvir-ritonavir prescription within +/-5 days. The few patients who received a prescription outside the +/-5 day treatment window were included in the untreated group. Patients who had more than one COVID-19 illness during the study period and therefore more than one potential index date were assigned the most recent index date.

PCC was defined by the presence of ≥1 ICD-10 diagnosis code for one or more of 45 new-onset symptoms or conditions, including the ICD-10 PCC code, U09.9, recorded ≥60 days after index date and prior to the end of follow-up on December 31, 2022 and not present 7-365 days prior to the index date. Due to the expansive definition of Long COVID, we relied on an extensive list of ICD-10 codes to ensure inclusivity and minimize the risk of overlooking potential PCCs (Supplemental Table 2).^22-25^

New symptoms and conditions were further grouped into 10 system categories. Patients qualified for each system category if they had at least 1 new-onset condition from that category ≥60 days after index date and had no other conditions from that category within 7-365 days prior to the index date.

Overall PCC was defined as the presence of ≥1 new-onset PCC symptom or condition and, separately, as ≥2 new-onset PCC symptoms or conditions. These patients may have had other symptoms or conditions within the year prior to index date.

### Statistical analysis

Treated and untreated patients were matched 1:2 on age, sex, month of index date, and HHS region.^26^ Person time was calculated from time of COVID-19 index date to date of new-onset PCC or end of follow-up. Analyses were executed using Databricks (v9.1 LTS, Databricks, Inc.) including Apache Spark (v3.1.2, Apache Software Foundation) and Python (v3.8.10, Python Software Foundation). Further analyses were conducted using R Statistical Software (v4.3.2, R Foundation 2023). Proportional hazard models and relative risk were calculated using the survival R package (v3.5-7, Therneau 2023); p-values <0.05 were considered significant.

We calculated incidence-based unadjusted relative risk (cumulative risk) to estimate the association between the nirmatrelvir-ritonavir and the overall measures of PCC (≥1 PCC, ≥2 PCC), 45 individual conditions, and 10 system categories (Supplemental Table 3). P-values (calculated using Fisher’s exact test) and 95% confidence intervals were produced using the fmsb R package (v0.7.6, Nakazawa 2023).

Proportional hazard models estimated the hazard ratios by age group (12-17, 18-49, 50-64, ≥65 years) for overall measures of PCC (≥1 PCC, ≥2 PCC), 45 individual conditions, and 10 system categories associated with nirmatrelvir-ritonavir in the main analysis. Proportionality assumption was tested using Schoenfeld residuals and visually examined survival curves. The survival curves did not cross in any cases, indicating that the nonproportionality might not lead to severe bias in the results.

Hazard ratios were adjusted for potential confounders identified a priori: age in years (continuous), sex, HHS region, obesity, number of underlying risk factors (1, 2, 3, 4, or ≥5), SARS-CoV-2 vaccination history (any vaccination ≥14 days prior to index date vs. no evidence of vaccination), and number of healthcare encounters in the year prior to index date.

We also performed several sensitivity and secondary analyses. To determine whether the analytic population was biased due to the inclusion of nirmatrelvir-ritonavir as a COVID-19 index event in the treated group, we limited the treated population to patients with a positive laboratory test for SARS CoV-2 or an ICD-10 diagnosis code.

To assess whether any apparent relationship between nirmatrelvir-ritonavir and PCC was in fact due to vaccination status – known to be under-reported in the dataset – we limited the analysis to persons with a record of at least one COVID-19 vaccination ≥14 days before index date. This model was adjusted for age group, sex, HHS region, obesity, categorical number of underlying risk factors, and number of healthcare encounters in the year prior to index date.

To assess the impact of underlying risk factors on the relationship between nirmatrelvir-ritonavir and PCC, we expanded the patient population to include patients <50 years with no underlying risk factors, who were excluded from the original analytic sample. We calculated incidence-based relative risk of overall PCC for persons aged 12-17, 18-49, and 50-64 years with no underlying risk factors and estimated hazard ratios of overall PCC for persons <50 years with no underlying risk factors. Treated and untreated patients were matched by age, sex, HHS region, and month of index date. Separately, we stratified the analysis by Charlson Comorbidity Index (CCI) score (0, 1-2, 3-4, ≥5), a weighted index using age and selected comorbid conditions to predict 10-year mortality.^27,28^ Patients were again matched by age, sex, HHS region, and month of index date.

This activity was reviewed by the CDC, deemed not research, and was conducted consistent with applicable federal law and CDC policy. (See e.g., 45 C.F.R. part 46.102(l)(2), 21 C.F.R. part 56; 42 U.S.C. §241(d); 5 U.S.C. §552a; 44 U.S.C. §3501 et seq.)

## Results

Patient enrollment and exclusions are shown in Figure 1. Over 4 million patients with COVID-19 during the study period met the inclusion criteria. Following exclusions, 291,433 treated patients were matched 1:2 to 582,866 untreated patients. Adults 50-64 years comprised the largest proportion of the analytic sample (44.1%); adolescents comprised 2.0% (Table 1). The sample was 62.0% female. Eleven percent of the sample had no underlying risk factors and 15.7% had ≥5. The most common underlying risk factors were mental health conditions (30.2%), heart conditions (25.0%), smoking (24.4%), and diabetes (23.6%). Nearly two-thirds of the sample was vaccinated (64.3%). The treated and untreated groups were similar across most characteristics, but a higher proportion of treated patients were vaccinated (69.8% vs. 61.6%), and a higher proportion of untreated patients had a prior COVID-19 infection >90 days before the index date (14.6% vs. 9.6%). Presence of underlying risk factors varied by age and the proportion of patients who had ≥ 5 underlying risk factors increased with age group (Supplement Table 4).

**Table 1.** Characteristics of COVID-19-positive patients treated or not treated with nirmatrelvir-ritonavir within ± 5 days of acute COVID-19 illness, HealthVerity, April 1-December 31, 2022.

|  | **Total** | **Treated<sup>a</sup>** | **Untreated<sup>b</sup>** |
| --- | --- | --- | --- |
|  | **N (%)** | **N (%)** | **N (%)** |
| **Total** | 874,299 (100.0) | 291,433 (100.0) | 582,866 (100.0) |
| **Age<sup>c</sup>** |  |  |  |
| 12-17 | 17,178 (2.0) | 5,726 (2.0) | 11,452 (2.0) |
| 18-49 | 292,818 (33.5) | 97,606 (33.5) | 195,212 (33.5) |
| 50-64 | 385,887 (44.1) | 128,629 (44.1) | 257,258 (44.1) |
| $\geq 65$ | 178,416 (20.4) | 59,472 (20.4) | 118,944 (20.4) |
| **Sex** |  |  |  |
| Female | 542,130 (62.0) | 180,710 (62.0) | 361,420 (62.0) |
| Male | 332,166 (38.0) | 110,722 (38.0) | 221,444 (38.0) |
| Missing/Unknown | 3 (0.0) | 1 (0.0) | 2 (0.0) |
| **Number of underlying risk factors** |  |  |  |
| 0 | 95,779 (11.0) | 33,623 (11.5) | 62,156 (10.7) |
| 1 | 219,918 (25.2) | 74,141 (25.4) | 145,777 (25.0) |
| 2 | 189,164 (21.6) | 64,290 (22.1) | 124,874 (21.4) |
| 3 | 139,039 (15.9) | 47,163 (16.2) | 91,876 (15.8) |
| 4 | 93,162 (10.7) | 30,764 (10.6) | 62,398 (10.7) |
| $\geq 5$ | 137,237 (15.7) | 41,452 (14.2) | 95,785 (16.4) |
| **Underlying risk factors** |  |  |  |
| Asthma | 155,984 (17.8) | 54,810 (18.8) | 101,174 (17.4) |
| Cancer | 66,503 (7.6) | 23,538 (8.1) | 42,965 (7.4) |
| Cerebrovascular disease | 60,851 (7.0) | 17,962 (6.2) | 42,889 (7.4) |
| Chronic kidney disease | 97,194 (11.1) | 28,769 (9.9) | 68,425 (11.7) |
| Chronic lung disease | 125,345 (14.3) | 38,734 (13.3) | 86,611 (14.9) |
| Chronic liver disease | 80,394 (9.2) | 27,287 (9.4) | 53,107 (9.1) |
| Cystic fibrosis | 274 (0.0) | 88 (0.0) | 186 (0.0) |
| Diabetes | 206,436 (23.6) | 67,707 (23.2) | 138,729 (23.8) |
| Disability | 140,451 (16.1) | 45,640 (15.7) | 94,811 (16.3) |
| Heart conditions | 218,530 (25.0) | 69,671 (23.9) | 148,859 (25.5) |
| HIV | 5,883 (0.7) | 2,082 (0.7) | 3,801 (0.7) |
| Mental health conditions | 263,658 (30.2) | 84,885 (29.1) | 178,773 (30.7) |
| Dementia | 12,771 (1.5) | 2,835 (1.0) | 9,936 (1.7) |
| Primary immunodeficiencies | 22,931 (2.6) | 8,463 (2.9) | 14,468 (2.5) |
| Smoking (current/former) | 213,729 (24.4) | 64,095 (22.0) | 149,634 (25.7) |
| Solid organ or blood stem cell transplant | 3,243 (0.4) | 924 (0.3) | 2,319 (0.4) |
| Tuberculosis | 1,165 (0.1) | 382 (0.1) | 783 (0.1) |
| Immunosuppressive medications | 1,895 (0.2) | 693 (0.2) | 1,202 (0.2) |
| **Obesity** |  |  |  |
| Yes | 358,371 (41.0) | 117,210 (40.2) | 241,161 (41.4) |
| No | 515,928 (59.0) | 174,223 (59.8) | 341,705 (58.6) |
| **Received $\geq 1$ COVID-19 vaccination 14+ days prior to index date** |  |  |  |
| Yes | 562,531 (64.3) | 203,341 (69.8) | 359,190 (61.6) |
| No | 311,768 (35.7) | 88,092 (30.2) | 223,676 (38.4) |
| **Prior COVID-19 infection (>90 days) before index date** |  |  |  |
| Yes | 112,967 (12.9) | 28,039 (9.6) | 84,928 (14.6) |
| No | 761,332 (87.1) | 263,394 (90.4) | 497,938 (85.4) |
| **Healthcare encounters in past year** |  |  |  |
| 0 | 23,964 (2.7) | 12,081 (4.1) | 11,883 (2.0) |
| 1-2 | 54,564 (6.2) | 17,002 (5.8) | 37,562 (6.4) |
| 3-5 | 114,570 (13.1) | 37,816 (13.0) | 76,754 (13.2) |
| 6-9 | 154,182 (17.6) | 51,834 (17.8) | 102,348 (17.6) |
| 10+ | 527,019 (60.3) | 172,700 (59.3) | 354,319 (60.8) |
| **HHS Region<sup>d</sup>** |  |  |  |
| Region 1 (CT, ME, MA, NH, RI, VT) | 19,647 (2.2) | 6,549 (2.2) | 13,098 (2.2) |
| Region 2 (NJ, NY, PR, VI) | 119,679 (13.7) | 39,893 (13.7) | 79,786 (13.7) |
| Region 3 (DE, DC, MD, PA, VA, WV) | 67,968 (7.8) | 22,656 (7.8) | 45,312 (7.8) |
| Region 4 (AL, FL, GA, KY, MS, NC, SC, TN) | 81,162 (9.3) | 27,054 (9.3) | 54,108 (9.3) |
| Region 5 (IL, IN, MI, MN, OH, WI) | 171,105 (19.6) | 57,035 (19.6) | 114,070 (19.6) |
| Region 6 (AR, LA, NM, OK, TX) | 183,243 (21.0) | 61,081 (21.0) | 122,162 (21.0) |
| Region 7 (IA, KS, MO, NE) | 20,511 (2.3) | 6,837 (2.3) | 13,674 (2.3) |
| Region 8 (CO, MT, ND, SD, UT, WY) | 13,197 (1.5) | 4,399 (1.5) | 8,798 (1.5) |
| Region 9 (AZ, CA, HI, NV, FM) | 180,918 (20.7) | 60,306 (20.7) | 120,612 (20.7) |
| Region 10 (AK, ID, OR, WA) | 16,650 (1.9) | 5,550 (1.9) | 11,100 (1.9) |
| Other | 219 (0.0) | 73 (0.0) | 146 (0.0) |
| **Charlson Comorbidity Index (CCI) Score<sup>e</sup>** |  |  |  |
| 0 | 201,346 (23.0) | 62,761 (21.5) | 138,585 (23.8) |
| 1-2 | 388,298 (44.4) | 135,247 (46.4) | 253,051 (43.4) |
| 3-4 | 172,065 (19.7) | 59,846 (20.5) | 112,219 (19.3) |
| 5+ | 112,590 (12.9) | 33,579 (11.5) | 79,011 (13.6) |

Abbreviations: HHS – U.S. Department of Health and Human Services
<sup>a</sup>Treated patients received a nirmatrelvir-ritonavir prescription within +/- 5 days of index date.
<sup>b</sup>Untreated patients did not receive a nirmatrelvir-ritonavir prescription within +/- 5 days of index date.
<sup>c</sup>All patients aged 12-17 and 18-49 years had one or more underlying risk factors in the main analysis.
<sup>d</sup>Health and Human Resources (HHS) regions are defined as follows:
Region 1: Connecticut, Maine, Massachusetts, New Hampshire, Rhode Island Vermont
Region 2: New Jersey, New York, Puerto Rico, US Virgin Islands
Region 3: Delaware, District of Columbia, Maryland, Pennsylvania, Virginia, West Virginia
Region 4: Alabama, Florida, Georgia, Kentucky, Mississippi, North Carolina, South Carolina, Tennessee
Region 5: Illinois, Indiana, Michigan, Minnesota, Ohio, Wisconsin
Region 6: Arkansas, Louisiana, New Mexico, Oklahoma, Texas
Region 7: Iowa, Kansas, Missouri, Nebraska
Region 8: Colorado, Montana, North Dakota, South Dakota, Utah, Wyoming
Region 9: Arizona, California, Hawaii, Nevada, American Samoa, Commonwealth of the Northern Mariana Islands, Federated States of Micronesia, Guam, Marshall Islands, and Republic of Palau

## Main analysis

Of 291,433 treated patients, 98,084 (33.7%) had ≥1 PCC and 40,147 (13.8%) had ≥2 PCCs during the study period; of 582,866 untreated patients, 208,033 (35.7%) had ≥1 PCC and 89,470 (15.4%) had ≥2 PCCs during the study period. The mean and median times to first PCC in the treated group were 111 days and 103 days, respectively, and 110 days and 101 days, respectively, in the untreated group.

### Unadjusted analysis

In an unadjusted analysis of incidence-based relative risk, the relationship between nirmatrelvir-ritonavir and PCC varied by age group: older adults aged 50-64 and ≥65 years had the most consistent reduction in risk of both overall PCC and individual symptoms and conditions. Younger adults aged 18-49 years had a small reduction in overall risk of PCC and a reduced risk of some but not all individual conditions; adolescents did not see any reduction in overall PCC or individual conditions associated with nirmatrelvir-ritonavir (Supplemental Table 5).

### Adjusted analysis

Among adults ≥65 years, nirmatrelvir-ritonavir was associated with a reduced risk of ≥1 symptom or condition (adjusted Hazard Ratio (aHR) 0.88, 95% CI 0.87-0.90) and ≥2 symptoms or conditions (aHR 0.82, 95% CI 0.80-0.84), as well as a reduced risk of all system category PCC and some individual conditions compared to untreated adults ≥65 years (Figure 2a; Supplemental Table 6).

**Figure 2.**
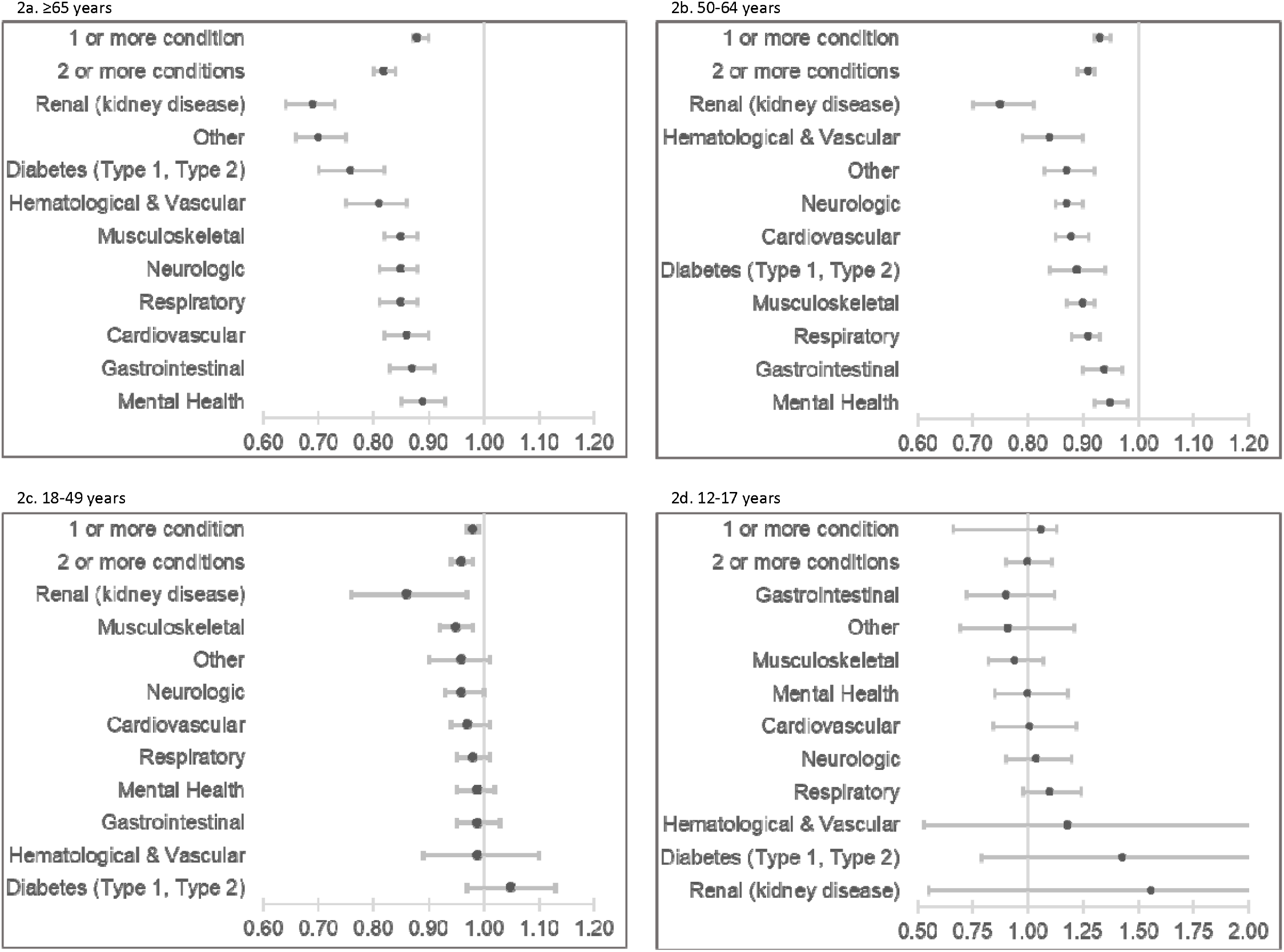
Adjusted hazard ratios and 95% confidence intervals of Post-COVID Conditions (PCC), overall and by system category, among persons treated versus untreated with nirmatrelvir-ritonavir during acute COVID-19 illlness, HealthVerity, April 1-December 31, 2022.

Among adults 50-64 years, nirmatrelvir-ritonavir treatment was associated with a reduced risk of ≥1 symptom or condition (aHR 0.93, 95% CI 0.92-0.95), ≥2 symptoms or conditions (aHR 0.91, 95% CI 0.89-0.92) and most individual and system symptoms and conditions (Figure 2b; Supplemental Table 6)

Among adults 18-49 years, nirmatrelvir-ritonavir treatment was associated with a reduced risk of ≥1 symptom or condition (aHR 0.98, 95% CI 0.97-0.99) and ≥2 symptoms or conditions (aHR 0.96, 95% CI 0.94-0.98) compared to untreated adults 18-49 years (Figure 2c; Supplemental Table 6). However, nirmatrelvir-ritonavir was associated with a reduced risk in fewer individual symptoms or conditions than in adults 50-64 and ≥65 years (Supplemental Table 6).

There was no association between nirmatrelvir-ritonavir treatment and overall PCC risk among adolescents, although there was an increased risk of hypertension and asthma among treated adolescents (Figure 2d; Supplemental Table 6).

## Secondary analyses

Limiting the analysis to patients with a positive laboratory test or ICD-10 diagnosis code as a COVID-19 index event resulted in slightly higher aHRs (Supplemental Table 7).

When analysis was limited to persons who received at least one SARS-CoV-2 vaccine ≥14 days before index event, nirmatrelvir-ritonavir treatment was associated with a reduced risk of ≥1 symptom or condition (aHR 0.94, 95% CI 0.93-0.95) and ≥2 symptoms or conditions (aHR 0.90, 95% CI 0.89-0.92) (Figure 3). In a sensitivity analysis limited to persons <65 years with no risk factors, nirmatrelvir-ritonavir treatment was associated with a reduced risk of overall PCC among adults aged 50-64 years (≥1 symptom or condition: aHR 0.90, 95% CI 0.87-0.93; ≥2 symptoms or conditions: aHR 0.88, 95% CD 0.84-0.93). Nirmatrelvir-ritonavir treatment was not associated with a change in PCC compared to untreated persons 12-17 and 18-49 years (Figure 3).

**Figure 3.**
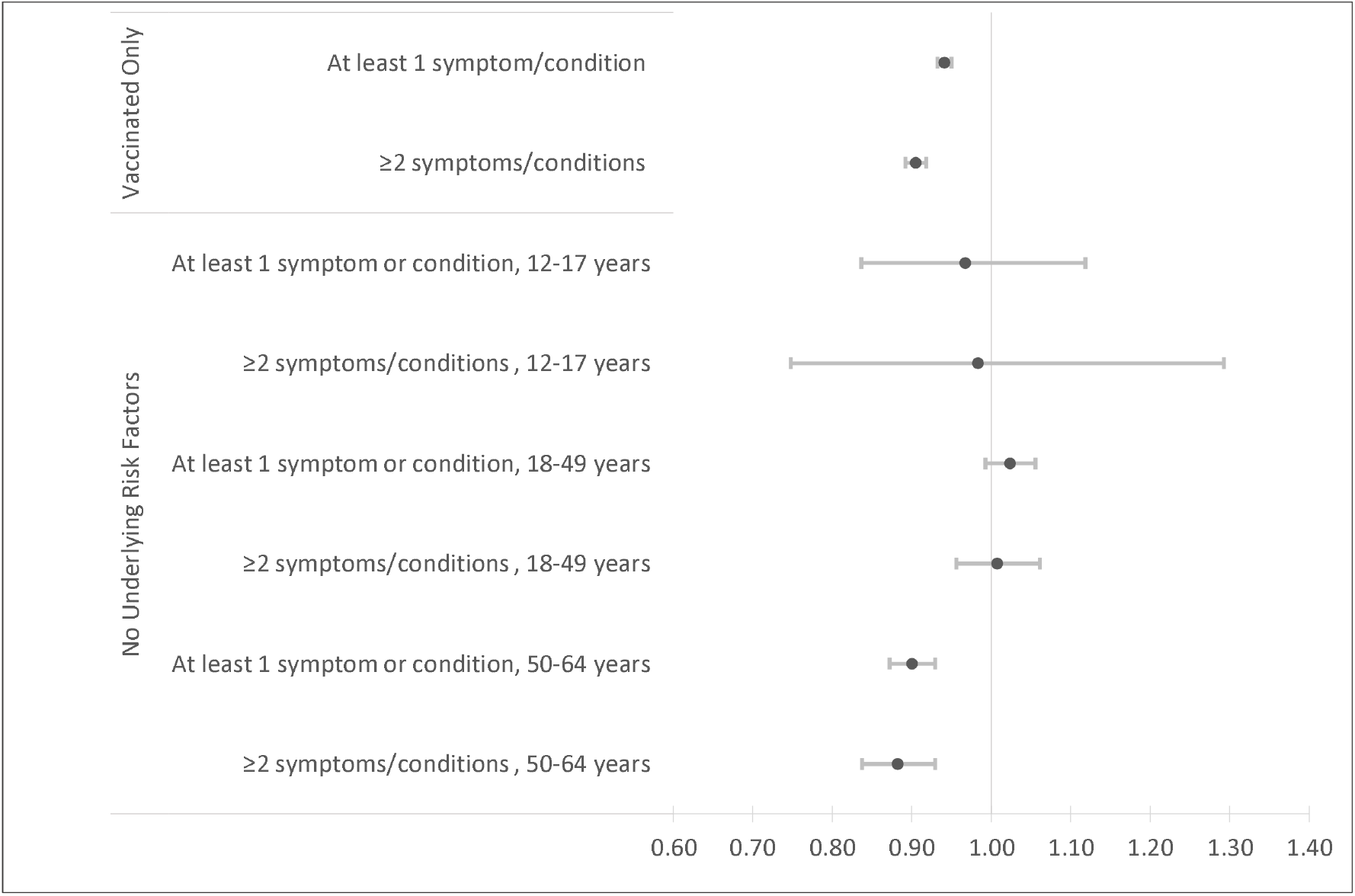
Adjusted hazard ratios of Post-COVID Conditions (PCC) among vaccinated^a,b^ patients (all ages) and among patients with no underlying risk factors^c^, by age group, treated versus untreated with nirmatrelvir-ritonavir during acute COVID-19 illness, HealthVerity, April 1-December 31, 2022. ^a^ Vaccination is defined as evidence of receipt of ≥1 COVID-19 vaccine ≥14 days before index date ^b^ Number of vaccinated patients: Treated=203,341; Untreated=359,160 ^c^ Number of patients with no underlying risk factors by age group: 12-17: Treated=1,466; Untreated=2,932 18-49: Treated=29,317; Untreated=58,634 50-64: Treated=23,398; Untreated=46,796

When stratified by CCI, there was a consistent relationship between nirmatrelvir-ritonavir treatment and lower relative risk of overall PCC among adults aged 50-64 years and ≥65 years, regardless of CCI score, but among adults aged 18-49 years with a CCI score of 0 there was no reduced risk. Treatment with nirmatrelvir-ritonavir was not associated with reduced risk of PCC in adolescents, regardless of CCI score (Table 2).

**Table 2.**
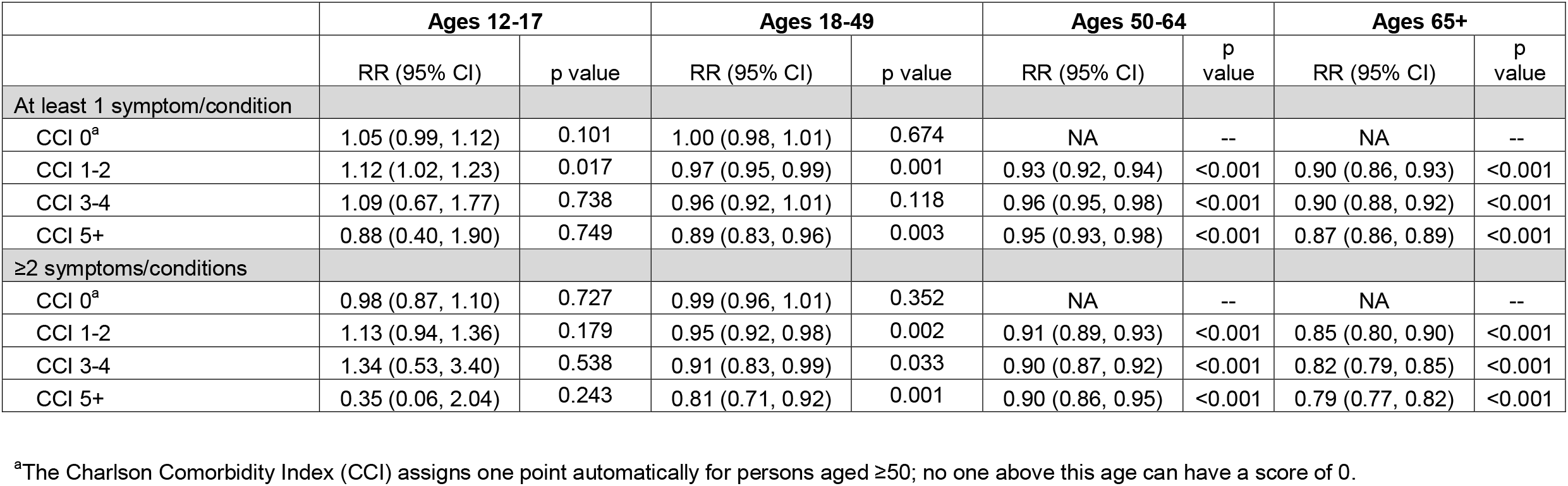
Incidence-based relative risk of at least 1 Post-COVID symptom or condition and at least 2 Post-COVID symptoms or conditions by Charlson Comorbidity Index (CCI) score, and age group among patients treated versus untreated with nirmatrelvir-ritonavir during acute COVID-19 illlness, HealthVerity, April 1-December 31, 2022.

|  | **Ages 12-17** |  | **Ages 18-49** |  | **Ages 50-64** |  | **Ages 65+** |  |
| --- | --- | --- | --- | --- | --- | --- | --- | --- |
|  | RR (95% CI) | p value | RR (95% CI) | p value | RR (95% CI) | p value | RR (95% CI) | p value |
| **At least 1 symptom/condition** |  |  |  |  |  |  |  |  |
| CCI 0<sup>a</sup> | 1.05 (0.99, 1.12) | 0.101 | 1.00 (0.98, 1.01) | 0.674 | NA | -- | NA | -- |
| CCI 1-2 | 1.12 (1.02, 1.23) | 0.017 | 0.97 (0.95, 0.99) | 0.001 | 0.93 (0.92, 0.94) | <0.001 | 0.90 (0.86, 0.93) | <0.001 |
| CCI 3-4 | 1.09 (0.67, 1.77) | 0.738 | 0.96 (0.92, 1.01) | 0.118 | 0.96 (0.95, 0.98) | <0.001 | 0.90 (0.88, 0.92) | <0.001 |
| CCI 5+ | 0.88 (0.40, 1.90) | 0.749 | 0.89 (0.83, 0.96) | 0.003 | 0.95 (0.93, 0.98) | <0.001 | 0.87 (0.86, 0.89) | <0.001 |
| **≥2 symptoms/conditions** |  |  |  |  |  |  |  |  |
| CCI 0<sup>a</sup> | 0.98 (0.87, 1.10) | 0.727 | 0.99 (0.96, 1.01) | 0.352 | NA | -- | NA | -- |
| CCI 1-2 | 1.13 (0.94, 1.36) | 0.179 | 0.95 (0.92, 0.98) | 0.002 | 0.91 (0.89, 0.93) | <0.001 | 0.85 (0.80, 0.90) | <0.001 |
| CCI 3-4 | 1.34 (0.53, 3.40) | 0.538 | 0.91 (0.83, 0.99) | 0.033 | 0.90 (0.87, 0.92) | <0.001 | 0.82 (0.79, 0.85) | <0.001 |
| CCI 5+ | 0.35 (0.06, 2.04) | 0.243 | 0.81 (0.71, 0.92) | 0.001 | 0.90 (0.86, 0.95) | <0.001 | 0.79 (0.77, 0.82) | <0.001 |

<sup>a</sup>The Charlson Comorbidity Index (CCI) assigns one point automatically for persons aged ≥50; no one above this age can have a score of 0.

## Discussion

Nirmatrelvir-ritonavir is recommended for use among persons ≥12 years with mild to moderate COVID-19 illness who are at risk for severe acute disease.^29^ This analysis suggests that non-hospitalized adults aged ≥50 years who take nirmatrelvir-ritonavir during the acute phase may additionally benefit from a reduced risk of PCC. There may be a slight reduction of risk of PCC among adults aged 18-49 years, although perhaps not clinically meaningful. There was no relationship between nirmatrelvir-ritonavir and overall PCC risk in high-risk adolescents, suggesting that nirmatrelvir-ritonavir may not be an effective tool for reducing the risk of PCC in adolescents.

Although nirmatrelvir-ritonavir was associated with reduced risk of PCC in adults 50-64 years with and without any underlying risk factors and regardless of CCI score, younger adults without underlying risk factors did not have a reduced risk. This supports treatment recommendations that suggest younger adults with certain underlying risk factors are more likely to benefit from treatment compared with younger adults without underlying conditions or risk factors.^29^

Nirmatrelvir-ritonavir’s benefits for reducing severe outcomes of acute illness have been well-studied,^5-11,30^ but there has been inconclusive evidence about whether it may also reduce the risk of PCC, and clinical trials have shown no impact of nirmatrelvir-ritonavir for *treatment* of PCC.^31,32^ This analysis utilizes a vast electronic healthcare dataset to capture valuable real world data about the relationship between nirmatrelvir-ritonavir and PCC as it is currently being used and focuses on people who have access to and seek medical care. This analysis corroborates findings from one observational study which showed a reduced risk of most individual symptoms and conditions among adults aged 60 years and older.^19^ However, our results run contrary to studies which showed no reduction in risk among person ≥18 years of age.^14,15^ Additionally, whereas some analyses have found a relationship only between nirmatrelvir-ritonavir and a reduced risk of specific conditions or symptoms (i.e., thromboembolic events,^16^ cognitive and fatigue symptoms clusters,^18^ or brain fog and chest pain^17^), our analysis found a relationship both overall and for numerous individual symptoms and conditions.

Strengths of our study include analysis of a large electronic healthcare records dataset that is representative of the healthcare-seeking population. Real-world data, such as that found in electronic healthcare records, provides insight on outcomes outside of the strict guidelines imposed by a clinical trial. Additionally, the analysis included adolescents and younger adults – populations that are affected by PCC but have been understudied. Moreover, the size of the dataset enabled us to stratify results by age and identify differential effects of nirmatrelvir-ritonavir in adolescents, younger adults, and older adults.

This analysis also had several limitations. First, although we can verify that a nirmatrelvir-ritonavir prescription was filled, we cannot verify adherence to treatment. However, non-adherence to treatment may bias the results towards the null, leading to more conservative estimates. Second, this was an observational study using electronic healthcare data and may have confounders beyond what was included in the models, such as prior SARS-CoV-2 infections or other factors that may influence nirmatrelvir-ritonavir prescriptions or the occurrence of PCC. For example, some models showed an increased risk of specific symptoms or conditions in treated patients. This could simply be due to chance, or it may be due to unmeasured confounding, such as confounding by indication, e.g., adolescents who received a prescription for nirmatrelvir-ritonavir differ in some way from those who did not receive a prescription. Additionally, misclassification bias might result due to variation in prescribing practices; we were unable to verify that clinicians were prescribing treatment per established guidelines. Third, a small number of patients who received treatment with nirmatrelvir-ritonavir outside the +/-5 day treatment window were included in the analysis, although this amounted to well under 1% in both the treated and untreated groups. Fourth, we used a broad definition of PCC and used the first occurrence of new-condition; moreover, and new conditions may not be due to SARS-CoV-2 infection. Fifth, the use of electronic healthcare data limits the analysis to persons who interacted with the healthcare system: persons who were eligible for nirmatrelvir-ritonavir but did not seek care were not captured in the data set. Similarly, patients who did not seek care related to PCC would be misclassified. Patients who had a history of a condition that was not captured in the 365 days prior to index date may also have been misclassified as having a new-onset PCC.

This observational study suggests that outpatient treatment of COVID-19 among older adults at high risk for severe disease might have an added benefit of reducing risk of PCC. Recent studies have found that nirmatrelvir-ritonavir is underutilized among older patients who are at risk of severe disease, despite the evidence of its benefits.^33-35^ Clinicians may consider treatment of persons with mild to moderate acute COVID-19 who are at high risk of severe disease, particularly those aged ≥50 years, not only to prevent hospitalization and death but also to mitigate possible long-term consequences.

## Conclusion

Overall, these results suggest a protective relationship between nirmatrelvir-ritonavir use during acute illness and risk of PCC among adults, particularly those aged ≥65 years, but not among adolescents aged 12-17 years. Consideration may be given to outpatient treatment of mild to moderate COVID-19 to reduce the risk of both severe disease and PCC.

## Data Availability

The data used in the present work are not publicly available.

## Notes

### Competing Interest Statement

The authors have declared no competing interest.

### Funding Statement

This project did not receive any funding.

### Author Declarations

The Human Research Protection Office of Centers for Disease Control and Prevention waived ethical approval for this work.

