## Supplemental Tables 3-7 for "Risk of Post-COVID Conditions among adolescents and adults who received nirmatrelvir-ritonavir for acute COVID-19: a retrospective cohort study"

Table of Contents

Supplemental Table 3. Post-COVID Conditions (PCC) symptoms and conditions included in analysis, by body system

Supplemental Table 4. Characteristics of COVID-19-positive patients treated or not treated with nirmatrelvir-ritonavir within ± 5 days of acute COVID-19 illlness, by age group, HealthVerity, April 1- December 31, 2022

Supplemental Table 5. Incidence-based unadjusted relative risk of overall Post-COVID Conditions (PCC), by system, and by individual symptom or condition by age group among patients treated with nirmatrelvir-ritonavir during acute COVID-19 illlness, HealthVerity, April 1- December 31, 2022

Supplemental Table 6. Adjusted hazard ratios for Post-COVID Conditions (PCC), by age group, among persons treated with nirmatrelvir-ritonavir during acute COVID-19 illness compared to untreated, Health Verity, April 1-December 31, 2022

Supplemental Table 7. Adjusted hazard ratios for Post-COVID Conditions (PCC) excluding patients with a nirmatrelvir-ritonavir COVID-19 index event, by age group, among persons treated with nirmatrelvir-ritonavir during acute COVID-19 illness compared to untreated, Health Verity, April 1-December 31, 2022

Supplemental Table 3. Post-COVID Conditions (PCC) symptoms and conditions included in analysis, by body system

**Overall PCC indicators**

At least 1 symptom/condition

≥2 symptoms/conditions

**Post-COVID-19 conditions**

**Cardiac**

Circulatory signs and symptoms

Coronary atherosclerosis and other heart disease

Heart Failure and chronic rheumatic heart disease

Hypertension

Cardiovascular disease

Cardiac dysrhythmia

Acute myocardial infarction

Myocarditis and cardiomyopathy

**Respiratory**

Respiratory symptoms and disease

Acute pulmonary embolism

Asthma

**Renal**

Kidney disease

**Hemolytic and vascular**

Coagulation and hemorrhagic event

Thromboembolic event

Cerebrovascular disease

**GI**

Change in Bowel Habits

Esophageal disorders

**Neurologic**

Ataxia / Trouble Walking

Autonomic Dysfunction

Cognitive disorders

Headache

Hearing loss and disturbances

Myoneural Disorders

Nervous System

Peripheral Nerve Disorders

Syncope and dizziness

Visual disturbances

Seizures

Sensations and Perceptions

Smell and taste disturbances

**Mental health**

Sleeping conditions

Other mental conditions

Substance-related disorder

Anxiety

Mood disorder

**Muscular**

Malaise and fatigue

Musculoskeletal pain

Muscle disorder

**Diabetes**

Diabetes Type 1

Diabetes Type 2

**Other**

Encephalitis

Weight Loss

Rash

PCC

Symptoms and signs involving cognition, perception, emotional state and behavior

Supplemental Table 4. Characteristics of COVID-19-positive patients treated or not treated with nirmatrelvir-ritonavir within ± 5 days of acute COVID-19 illness, by age group, HealthVerity, April 1- December 31, 2022

|  | **12-17 years** | | **18-49 years** | | **50-64 years** | | **≥65 years** | |
| --- | --- | --- | --- | --- | --- | --- | --- | --- |
|  | **Paxlovid N (%)** | **No Paxlovid N (%)** | **Paxlovid N (%)** | **No Paxlovid N (%)** | **Paxlovid N (%)** | **No Paxlovid N (%)** | **Paxlovid N (%)** | **No Paxlovid N (%)** |
| **Total** | 5,726 (100.0) | 11,452 (100.0) | 97,606 (100.0) | 195,212 (100.0) | 128,629 (100.0) | 257,258 (100.0) | 59,472 (100.0) | 118,944 (100.0) |
| **Sex** |  |  |  |  |  |  |  |  |
| Female | 2,939 (51.3) | 5,878 (51.3) | 64,240 (65.8) | 128,480 (65.8) | 78,840 (61.3) | 157,680 (61.3) | 34,691 (58.3) | 69,382 (58.3) |
| Male | 2,787 (48.7) | 5,574 (48.7) | 33,366 (34.2) | 66,732 (34.2) | 49,788 (38.7) | 99,576 (38.7) | 24,781 (41.7) | 49,562 (41.7) |
| Missing/Unknown |  |  |  |  | 1 (0.0) | 2 (0.0) |  |  |
| **Number of underlying conditions** |  |  |  |  |  |  |  |  |
| 0 | 0 (0.0) | 0 (0.0) | 0 (0.0) | 0 (0.0) | 24,579 (19.1) | 48,696 (18.9) | 9,044 (15.2) | 13,460 (11.3) |
| 1 | 2,014 (35.2) | 4,638 (40.5) | 31,333 (32.1) | 67,137 (34.4) | 29,255 (22.7) | 55,690 (21.6) | 11,539 (19.4) | 18,312 (15.4) |
| 2 | 1,697 (29.6) | 3,634 (31.7) | 25,695 (26.3) | 52,324 (26.8) | 25,767 (20.0) | 49,090 (19.1) | 11,131 (18.7) | 19,826 (16.7) |
| 3 | 1,073 (18.7) | 1,940 (16.9) | 17,820 (18.3) | 34,058 (17.4) | 19,132 (14.9) | 37,549 (14.6) | 9,138 (15.4) | 18,329 (15.4) |
| 4 | 562 (9.8) | 852 (7.4) | 10,711 (11.0) | 20,017 (10.3) | 12,589 (9.8) | 26,173 (10.2) | 6,902 (11.6) | 15,356 (12.9) |
| ≥5 | 380 (6.6) | 388 (3.4) | 12,047 (12.3) | 21,676 (11.1) | 17,307 (13.5) | 40,060 (15.6) | 11,718 (19.7) | 33,661 (28.3) |
| **Underlying medical conditions** |  |  |  |  |  |  |  |  |
| Asthma | 2,042 (35.7) | 3,133 (27.4) | 24,809 (25.4) | 41,327 (21.2) | 20,709 (16.1) | 40,060 (15.6) | 7,250 (12.2) | 16,654 (14.0) |
| Cancer | 79 (1.4) | 68 (0.6) | 3,930 (4.0) | 6,006 (3.1) | 11,235 (8.7) | 19,987 (7.8) | 8,294 (13.9) | 16,904 (14.2) |
| Cerebrovascular disease | 30 (0.5) | 44 (0.4) | 2,316 (2.4) | 4,556 (2.3) | 7,729 (6.0) | 17,464 (6.8) | 7,887 (13.3) | 20,825 (17.5) |
| Chronic kidney disease | 135 (2.4) | 158 (1.4) | 6,369 (6.5) | 12,629 (6.5) | 11,174 (8.7) | 26,386 (10.3) | 11,091 (18.6) | 29,252 (24.6) |
| Chronic lung disease | 303 (5.3) | 500 (4.4) | 10,629 (10.9) | 20,175 (10.3) | 17,071 (13.3) | 38,108 (14.8) | 10,731 (18.0) | 27,828 (23.4) |
| Chronic liver disease | 164 (2.9) | 209 (1.8) | 9,925 (10.2) | 17,526 (9.0) | 13,047 (10.1) | 26,248 (10.2) | 4,151 (7.0) | 9,124 (7.7) |
| Cystic fibrosis | 7 (0.1) | 9 (0.1) | 50 (0.1) | 72 (0.0) | 18 (0.0) | 79 (0.0) | 13 (0.0) | 26 (0.0) |
| Diabetes | 254 (4.4) | 263 (2.3) | 16,585 (17.0) | 28,042 (14.4) | 31,693 (24.6) | 65,169 (25.3) | 19,175 (32.2) | 45,255 (38.0) |
| Disability | 2,153 (37.6) | 3,923 (34.3) | 20,630 (21.1) | 37,590 (19.3) | 15,321 (11.9) | 31,970 (12.4) | 7,536 (12.7) | 21,328 (17.9) |
| Heart conditions | 383 (6.7) | 584 (5.1) | 14,650 (15.0) | 27,508 (14.1) | 31,279 (24.3) | 65,145 (25.3) | 23,359 (39.3) | 55,622 (46.8) |
| HIV | 06 (0.1) | 04 (0.0) | 931 (1.0) | 1,491 (0.8) | 971 (0.8) | 1,792 (0.7) | 174 (0.3) | 514 (0.4) |
| Mental health conditions | 1,621 (28.3) | 3,181 (27.8) | 39,539 (40.5) | 77,557 (39.7) | 31,733 (24.7) | 67,384 (26.2) | 11,992 (20.2) | 30,651 (25.8) |
| Dementia | 0 (0.0) | 2 (0.0) | 139 (0.1) | 276 (0.1) | 390 (0.3) | 1,207 (0.5) | 2,306 (3.9) | 8,451 (7.1) |
| Primary immunodeficiencies | 175 (3.1) | 121 (1.1) | 2,878 (2.9) | 3,879 (2.0) | 3,757 (2.9) | 6,852 (2.7) | 1,653 (2.8) | 3,616 (3.0) |
| Smoking (current/former) | 82 (1.4) | 192 (1.7) | 24,181 (24.8) | 57,458 (29.4) | 26,883 (20.9) | 61,331 (23.8) | 12,949 (21.8) | 30,653 (25.8) |
| Solid organ or blood stem cell transplant | 18 (0.3) | 9 (0.1) | 268 (0.3) | 524 (0.3) | 399 (0.3) | 1,088 (0.4) | 239 (0.4) | 698 (0.6) |
| Tuberculosis | 6 (0.1) | 7 (0.1) | 150 (0.2) | 275 (0.1) | 152 (0.1) | 328 (0.1) | 74 (0.1) | 173 (0.1) |
| Immunosuppressive medications | 3 (0.1) | 4 (0.0) | 94 (0.1) | 121 (0.1) | 336 (0.3) | 516 (0.2) | 260 (0.4) | 561 (0.5) |
| **Obesity** |  |  |  |  |  |  |  |  |
| Yes | 2,283 (39.9) | 4,017 (35.1) | 47,858 (49.0) | 93,085 (47.7) | 48,604 (37.8) | 103,172 (40.1) | 18,465 (31.0) | 40,887 (34.4) |
| No | 3,443 (60.1) | 7,435 (64.9) | 49,748 (51.0) | 102,127 (52.3) | 80,025 (62.2) | 154,086 (59.9) | 41,007 (69.0) | 78,057 (65.6) |
| **Received COVID-19 vaccination 14+ days prior** |  |  |  |  |  |  |  |  |
| Yes | 3,129 (54.6) | 5,122 (44.7) | 63,017 (64.6) | 107,223 (54.9) | 96,239 (74.8) | 173,133 (67.3) | 40,956 (68.9) | 73,712 (62.0) |
| No | 2,597 (45.4) | 6,330 (55.3) | 34,589 (35.4) | 87,989 (45.1) | 32,390 (25.2) | 84,125 (32.7) | 18,516 (31.1) | 45,232 (38.0) |
| **Prior COVID-19 infection (>90 days)?** |  |  |  |  |  |  |  |  |
| Yes | 690 (12.1) | 1,793 (15.7) | 13,395 (13.7) | 36,237 (18.6) | 11,131 (8.7) | 34,942 (13.6) | 2,823 (4.7) | 11,956 (10.1) |
| No | 5,036 (87.9) | 9,659 (84.3) | 84,211 (86.3) | 158,975 (81.4) | 117,498 (91.3) | 222,316 (86.4) | 56,649 (95.3) | 106,988 (89.9) |
| **Healthcare encounters in past year** |  |  |  |  |  |  |  |  |
| 0 | 151 (2.6) | 215 (1.9) | 3,626 (3.7) | 4,806 (2.5) | 7,351 (5.7) | 5,622 (2.2) | 953 (1.6) | 1,240 (1.0) |
| 1-2 | 493 (8.6) | 1,178 (10.3) | 5,823 (6.0) | 14,565 (7.5) | 7,005 (5.4) | 15,741 (6.1) | 3,681 (6.2) | 6,078 (5.1) |
| 3-5 | 1,068 (18.7) | 2,460 (21.5) | 12,983 (13.3) | 28,870 (14.8) | 15,692 (12.2) | 32,729 (12.7) | 8,073 (13.6) | 12,695 (10.7) |
| 6-9 | 1,184 (20.7) | 2,626 (22.9) | 16,990 (17.4) | 36,600 (18.7) | 22,967 (17.9) | 45,616 (17.7) | 10,693 (18.0) | 17,506 (14.7) |
| 10+ | 2,830 (49.4) | 4,973 (43.4) | 58,184 (59.6) | 110,371 (56.5) | 75,614 (58.8) | 157,550 (61.2) | 36,072 (60.7) | 81,425 (68.5) |
| **HHS Region** |  |  |  |  |  |  |  |  |
| Region 1 (CT, ME, MA, NH, RI, VT) | 91 (1.6) | 182 (1.6) | 2,322 (2.4) | 4,644 (2.4) | 3,068 (2.4) | 6,136 (2.4) | 1,068 (1.8) | 2,136 (1.8) |
| Region 2 (NJ, NY, PR, VI) | 374 (6.5) | 748 (6.5) | 10,961 (11.2) | 21,922 (11.2) | 18,374 (14.3) | 36,748 (14.3) | 10,184 (17.1) | 20,368 (17.1) |
| Region 3 (DE, DC, MD, PA, VA, WV) | 291 (5.1) | 582 (5.1) | 6,647 (6.8) | 13,294 (6.8) | 8,845 (6.9) | 17,690 (6.9) | 6,873 (11.6) | 13,746 (11.6) |
| Region 4 (AL, FL, GA, KY, MS, NC, SC, TN) | 1,011 (17.7) | 2,022 (17.7) | 9,771 (10.0) | 19,542 (10.0) | 9,901 (7.7) | 19,802 (7.7) | 6,371 (10.7) | 12,742 (10.7) |
| Region 5 (IL, IN, MI, MN, OH, WI) | 592 (10.3) | 1,184 (10.3) | 18,804 (19.3) | 37,608 (19.3) | 26,712 (20.8) | 53,424 (20.8) | 10,927 (18.4) | 21,854 (18.4) |
| Region 6 (AR, LA, NM, OK, TX) | 1,817 (31.7) | 3,634 (31.7) | 23,442 (24.0) | 46,884 (24.0) | 27,295 (21.2) | 54,590 (21.2) | 8,527 (14.3) | 17,054 (14.3) |
| Region 7 (IA, KS, MO, NE) | 137 (2.4) | 274 (2.4) | 2,314 (2.4) | 4,628 (2.4) | 2,266 (1.8) | 4,532 (1.8) | 2,120 (3.6) | 4,240 (3.6) |
| Region 8 (CO, MT, ND, SD, UT, WY) | 44 (0.8) | 88 (0.8) | 1,152 (1.2) | 2,304 (1.2) | 1,955 (1.5) | 3,910 (1.5) | 1,248 (2.1) | 2,496 (2.1) |
| Region 9 (AZ, CA, HI, NV, FM) | 1,328 (23.2) | 2,656 (23.2) | 20,839 (21.4) | 41,678 (21.4) | 27,885 (21.7) | 55,770 (21.7) | 10,254 (17.2) | 20,508 (17.2) |
| Region 10 (AK, ID, OR, WA) | 41 (0.7) | 82 (0.7) | 1,346 (1.4) | 2,692 (1.4) | 2,288 (1.8) | 4,576 (1.8) | 1,875 (3.2) | 3,750 (3.2) |
| Other | 0 (0.0) | 0 (0.0) | 8 (0.0) | 16 (0.0) | 40 (0.0) | 80 (0.0) | 25 (0.0) | 50 (0.0) |

Supplemental Table 5. Incidence-based unadjusted relative risk of overall Post-COVID Conditions (PCC), by system, and by individual symptom or condition by age group among patients treated with nirmatrelvir-ritonavir during acute COVID-19 illness, HealthVerity, April 1- December 31, 2022.

|  | **Ages 12-17** | | **Ages 18-49** | | **Ages 50-64** | | **Ages 65+** | |
| --- | --- | --- | --- | --- | --- | --- | --- | --- |
|  | **RR (95% CI)** | **p value** | **RR (95% CI)** | **p value** | **RR (95% CI)** | **p value** | **RR (95% CI)** | **p value** |
| **Experienced post-COVID-19 symptoms/conditions?** |  |  |  |  |  |  |  |  |
| At least 1 symptom/condition | 1.04 (0.99, 1.09) | >0.05 | 0.99 (0.98, 1.00) | <0.05 | 0.95 (0.94, 0.96) | <0.01 | 0.91 (0.90, 0.92) | <0.01 |
| ≥2 symptoms/conditions | 1.03 (0.94, 1.13) | >0.05 | 0.96 (0.94, 0.98) | <0.01 | 0.92 (0.90, 0.93) | <0.01 | 0.84 (0.82, 0.86) | <0.01 |
| **Post-COVID Conditions** |  |  |  |  |  |  |  |  |
| **Cardiovascular** | **1.08 (0.91, 1.29)** | **>0.05** | **1.0 (1.0, 1.0)** | **>0.05** | **0.89 (0.87, 0.92)** | **<0.01** | **0.86 (0.82, 0.90)** | **<0.01** |
| Circulatory signs and symptoms | 1.14 (0.95, 1.38) | >0.05 | 0.95 (0.92, 0.99) | <0.01 | 0.91 (0.88, 0.94) | <0.01 | 0.88 (0.83, 0.92) | <0.01 |
| Coronary atherosclerosis and other heart disease | 0.00 (0.00, 0.00) | 0 | 0.94 (0.83, 1.05) | >0.05 | 0.92 (0.87, 0.97) | <0.01 | 0.82 (0.76, 0.87) | <0.01 |
| Heart Failure and chronic rheumatic heart disease | 2.00 (0.28, 14.21) | >0.05 | 0.97 (0.79, 1.18) | >0.05 | 0.78 (0.69, 0.89) | <0.01 | 0.85 (0.75, 0.96) | <0.05 |
| Hypertension | 1.33 (0.87, 2.04) | >0.05 | 0.99 (0.95, 1.03) | >0.05 | 0.88 (0.85, 0.91) | <0.01 | 0.83 (0.79, 0.88) | <0.01 |
| Cardiovascular disease | 0.00 (0.00, 0.00) | 0 | 0.64 (0.27, 1.49) | >0.05 | 1.05 (0.68, 1.62) | >0.05 | 0.67 (0.39, 1.15) | >0.05 |
| Cardiac dysrhythmia | 0.95 (0.59, 1.51) | >0.05 | 0.93 (0.85, 1.01) | >0.05 | 0.91 (0.85, 0.96) | <0.01 | 0.80 (0.75, 0.86) | <0.01 |
| Acute myocardial infarction | 0.00 (0.00, 0.00) | 0 | 0.83 (0.65, 1.08) | >0.05 | 0.80 (0.70, 0.92) | <0.01 | 0.70 (0.60, 0.81) | <0.01 |
| Myocarditis and cardiomyopathy | 2.67 (0.60, 11.92) | >0.05 | 1.10 (0.90, 1.36) | >0.05 | 0.81 (0.70, 0.93) | <0.01 | 0.72 (0.62, 0.83) | <0.01 |
| **Respiratory** | **1.05 (0.95, 1.16)** | **>0.05** | **0.98 (0.95, 1.01)** | **>0.05** | **0.92 (0.90, 0.94)** | **<0.01** | **0.87 (0.83, 0.90)** | **<0.01** |
| Respiratory symptoms and disease | 1.06 (0.96, 1.17) | >0.05 | 0.95 (0.93, 0.98) | <0.01 | 0.91 (0.89, 0.94) | <0.01 | 0.86 (0.83, 0.90) | <0.01 |
| Acute pulmonary embolism | 2.00 (0.13, 31.97) | >0.05 | 0.71 (0.55, 0.91) | <0.01 | 0.79 (0.66, 0.94) | <0.01 | 0.71 (0.57, 0.87) | <0.01 |
| Asthma | 1.24 (1.04, 1.48) | <0.05 | 1.17 (1.12, 1.23) | <0.01 | 1.01 (0.95, 1.06) | >0.05 | 0.87 (0.79, 0.95) | <0.01 |
| **Renal** |  |  |  |  |  |  |  |  |
| Kidney disease | 1.34 (0.48, 3.75) | >0.05 | 0.81 (0.72, 0.91) | <0.01 | 0.75 (0.70, 0.80) | <0.01 | 0.70 (0.65, 0.75) | <0.01 |
| **Hemolytic and vascular** | **0.91 (0.43, 1.93)** | **>0.05** | **0.96 (0.87, 1.06)** | **>0.05** | **0.83 (0.77, 0.89)** | **<0.01** | **0.78 (0.72, 0.83)** | **<0.01** |
| Coagulation and hemorrhagic disorders | 0.90 (0.41, 1.98) | >0.05 | 1.00 (0.89, 1.11) | >0.05 | 0.88 (0.80, 0.96) | <0.01 | 0.81 (0.74, 0.89) | <0.01 |
| Thromboembolic event | 0.00 (0.00, 0.00) | 0 | 0.79 (0.63, 0.99) | <0.05 | 0.62 (0.53, 0.74) | <0.01 | 0.69 (0.57, 0.82) | <0.01 |
| Cerebrovascular disease | 1.00 (0.09, 11.03) | >0.05 | 0.86 (0.68, 1.10) | >0.05 | 0.81 (0.71, 0.92) | <0.01 | 0.73 (0.65, 0.83) | <0.01 |
| **Gastrointestinal** | **0.87 (0.71, 1.07)** | **>0.05** | **0.97 (0.94, 1.01)** | **>0.05** | **0.94 (0.91, 0.97)** | **<0.01** | **0.86 (0.82, 0.90)** | **<0.01** |
| Change in Bowel Habits | 0.89 (0.70, 1.13) | >0.05 | 0.98 (0.93, 1.03) | >0.05 | 0.90 (0.86, 0.94) | <0.01 | 0.84 (0.78, 0.89) | <0.01 |
| Esophageal disorders | 0.88 (0.65, 1.17) | >0.05 | 0.98 (0.94, 1.02) | >0.05 | 0.95 (0.91, 0.98) | <0.01 | 0.87 (0.83, 0.92) | <0.01 |
| **Neurologic** | **1.11 (0.98, 1.26)** | **>0.05** | **0.96 (0.93, 0.98)** | **<0.01** | **0.88 (0.85, 0.90)** | **<0.01** | **0.88 (0.84, 0.91)** | **<0.01** |
| Ataxia / Trouble Walking | 1.16 (0.74, 1.80) | >0.05 | 1.04 (0.96, 1.14) | >0.05 | 0.80 (0.75, 0.86) | <0.01 | 0.74 (0.69, 0.79) | <0.01 |
| Autonomic Dysfunction | 2.01 (0.65, 6.22) | >0.05 | 1.04 (0.85, 1.27) | >0.05 | 0.89 (0.73, 1.07) | >0.05 | 0.89 (0.68, 1.15) | >0.05 |
| Cognitive disorders | 1.25 (0.41, 3.82) | >0.05 | 0.83 (0.69, 1.00) | <0.05 | 0.85 (0.71, 1.01) | >0.05 | 0.63 (0.50, 0.80) | <0.01 |
| Headache | 1.04 (0.90, 1.21) | >0.05 | 0.92 (0.89, 0.96) | <0.01 | 0.89 (0.86, 0.93) | <0.01 | 0.89 (0.83, 0.96) | <0.01 |
| Hearing loss and disturbances | 1.27 (0.78, 2.07) | >0.05 | 1.19 (1.07, 1.33) | <0.01 | 0.95 (0.87, 1.03) | >0.05 | 1.07 (0.96, 1.18) | >0.05 |
| Myoneural Disorders | 3.34 (0.80, 13.96) | >0.05 | 1.14 (0.92, 1.42) | >0.05 | 1.00 (0.84, 1.19) | >0.05 | 0.83 (0.63, 1.11) | >0.05 |
| Nervous System | 1.43 (0.45, 4.51) | >0.05 | 0.87 (0.72, 1.04) | >0.05 | 0.89 (0.74, 1.07) | >0.05 | 0.65 (0.47, 0.90) | <0.01 |
| Peripheral Nerve Disorders | 1.20 (0.59, 2.46) | >0.05 | 0.96 (0.90, 1.02) | >0.05 | 0.93 (0.89, 0.98) | <0.01 | 0.90 (0.83, 0.97) | <0.01 |
| Syncope and dizziness | 1.00 (0.81, 1.24) | >0.05 | 0.92 (0.87, 0.96) | <0.01 | 0.88 (0.84, 0.92) | <0.01 | 0.86 (0.81, 0.91) | <0.01 |
| Visual disturbances | 1.43 (0.90, 2.29) | >0.05 | 0.98 (0.88, 1.10) | >0.05 | 0.96 (0.86, 1.07) | >0.05 | 1.03 (0.89, 1.20) | >0.05 |
| Seizures | 1.16 (0.61, 2.19) | >0.05 | 0.85 (0.72, 1.00) | <0.05 | 0.66 (0.54, 0.80) | <0.01 | 0.66 (0.51, 0.84) | <0.01 |
| Sensations and Perceptions | 0.00 (0.00, 0.00) | 0 | 0.70 (0.52, 0.95) | <0.05 | 0.71 (0.50, 1.00) | <0.05 | 0.86 (0.57, 1.29) | >0.05 |
| Smell and taste disturbances | 3.99 (0.36, 44.05) | >0.05 | 0.98 (0.73, 1.33) | >0.05 | 1.00 (0.73, 1.36) | >0.05 | 1.02 (0.63, 1.65) | >0.05 |
| **Mental health** | **0.98 (0.85, 1.14)** | **>0.05** | **1.01 (0.98, 1.04)** | **>0.05** | **0.95 (0.92, 0.98)** | **<0.01** | **0.88 (0.84, 0.93)** | **<0.01** |
| Sleeping conditions | 0.87 (0.67, 1.14) | >0.05 | 1.04 (0.99, 1.08) | >0.05 | 1.00 (0.97, 1.04) | >0.05 | 0.91 (0.85, 0.96) | <0.01 |
| Other mental conditions | 0.75 (0.54, 1.06) | >0.05 | 0.80 (0.71, 0.91) | <0.01 | 0.76 (0.66, 0.88) | <0.01 | 0.63 (0.51, 0.78) | <0.01 |
| Substance-related disorder | 0.48 (0.21, 1.10) | >0.05 | 0.59 (0.52, 0.66) | <0.01 | 0.65 (0.57, 0.75) | <0.01 | 0.63 (0.50, 0.78) | <0.01 |
| Anxiety | 1.08 (0.92, 1.26) | >0.05 | 1.00 (0.97, 1.04) | >0.05 | 0.93 (0.89, 0.96) | <0.01 | 0.89 (0.83, 0.94) | <0.01 |
| Mood disorder | 0.93 (0.76, 1.14) | >0.05 | 0.92 (0.88, 0.96) | <0.01 | 0.93 (0.88, 0.99) | <0.05 | 0.82 (0.75, 0.89) | <0.01 |
| **Musculoskeletal** | **0.96 (0.86, 1.08)** | **>0.05** | **0.97 (0.94, 0.99)** | **<0.01** | **0.90 (0.88, 0.92)** | **<0.01** | **0.87 (0.84, 0.91)** | **<0.01** |
| Malaise and fatigue | 1.09 (0.91, 1.31) | >0.05 | 1.01 (0.98, 1.05) | >0.05 | 0.93 (0.90, 0.96) | <0.01 | 0.83 (0.79, 0.88) | <0.01 |
| Musculoskeletal pain | 0.97 (0.86, 1.10) | >0.05 | 0.96 (0.93, 0.98) | <0.01 | 0.91 (0.89, 0.94) | <0.01 | 0.91 (0.88, 0.95) | <0.01 |
| Muscle disorder | 0.82 (0.54, 1.25) | >0.05 | 0.97 (0.91, 1.03) | >0.05 | 0.86 (0.82, 0.91) | <0.01 | 0.64 (0.59, 0.69) | <0.01 |
| **Diabetes** | **1.28 (0.72, 2.28)** | **>0.05** | **1.04 (0.97, 1.11)** | **>0.05** | **0.88 (0.83, 0.93)** | **<0.01** | **0.74 (0.68, 0.80)** | **<0.01** |
| Diabetes Type 1 | 0.87 (0.22, 3.35) | >0.05 | 1.09 (0.85, 1.40) | >0.05 | 0.94 (0.78, 1.14) | >0.05 | 0.67 (0.51, 0.88) | <0.01 |
| Diabetes Type 2 | 1.52 (0.88, 2.65) | >0.05 | 1.06 (0.99, 1.13) | >0.05 | 0.90 (0.85, 0.95) | <0.01 | 0.74 (0.68, 0.80) | <0.01 |
| **Other** | **0.89 (0.69, 1.15)** | **>0.05** | **0.95 (0.90, 1.01)** | **>0.05** | **0.87 (0.83, 0.91)** | **<0.01** | **0.70 (0.66, 0.74)** | **<0.01** |
| Encephalitis | 0.00 (0.00, 0.00) | 0 | 0.00 (0.00, 0.00) | 0 | 0.29 (0.04, 2.32) | >0.05 | 0.67 (0.07, 6.41) | >0.05 |
| Weight Loss | 0.90 (0.63, 1.29) | >0.05 | 0.86 (0.77, 0.95) | <0.01 | 0.83 (0.76, 0.91) | <0.01 | 0.64 (0.58, 0.71) | <0.01 |
| Rash | 0.31 (0.07, 1.36) | >0.05 | 0.99 (0.84, 1.17) | >0.05 | 1.12 (0.96, 1.31) | >0.05 | 0.94 (0.67, 1.32) | >0.05 |
| PCC | 2.62 (1.27, 5.40) | <0.01 | 1.04 (0.94, 1.15) | >0.05 | 0.86 (0.79, 0.94) | <0.01 | 0.68 (0.60, 0.78) | <0.01 |
| Symptoms and signs involving cognition, perception, emotional state and behavior | 0.78 (0.50, 1.21) | >0.05 | 0.91 (0.84, 0.99) | <0.05 | 0.81 (0.76, 0.88) | <0.01 | 0.67 (0.62, 0.73) | <0.01 |

Supplemental Table 6. Adjusted^a^ hazard ratios for Post-COVID Conditions (PCC), by age group, among persons treated with nirmatrelvir-ritonavir during acute COVID-19 illness compared to untreated, Health Verity, April 1-December 31, 2022.

|  | **Overall (All ages)** | | **Ages 12-17** | | | **Ages 18-49** | | | **Ages 50-64** | | | **Ages 65+** | |
| --- | --- | --- | --- | --- | --- | --- | --- | --- | --- | --- | --- | --- | --- |
|  | **Hazard Ratio**  **(95% CI)** | **p** | **Hazard Ratio**  **(95% CI)** | | **p** | **Hazard Ratio**  **(95% CI)** | **p** | | **Hazard Ratio**  **(95% CI)** | **p** | | **Hazard Ratio**  **(95% CI)** | **p** |
| **Experienced Post-COVID Conditions** |  | |  |  | |  | |  |  | |  |  | |
| At least 1 symptom/condition | 0.94 (0.93, 0.95) | <0.01 | 1.06 (0.66, 1.13) | | >0.05 | 0.98 (0.97, 0.99) | <0.01 | | 0.93 (0.92, 0.95) | <0.01 | | 0.88 (0.87, 0.90) | <0.01 |
| ≥2 symptoms/conditions | 0.91 (0.90, 0.92) | <0.01 | 1.00 (0.90, 1.11) | | >0.05 | 0.96 (0.94, 0.98) | <0.01 | | 0.91 (0.89, 0.92) | <0.01 | | 0.82 (0.80, 0.84) | <0.01 |
| **Post-COVID Conditions** |  |  |  | |  |  |  | |  |  | |  |  |
| **Cardiovascular** | **0.91 (0.90, 0.93)** | **<0.01** | **1.01 (0.84, 1.22)** | | **>0.05** | **0.97 (0.94, 1.01)** | **>0.05** | | **0.88 (0.85, 0.91)** | **<0.01** | | **0.86 (0.82, 0.90)** | **<0.01** |
| Circulatory signs and symptoms | 0.92 (0.90, 0.95) | <0.01 | 0.99 (0.81, 1.21) | | >0.05 | 0.96 (0.93, 1.00) | >0.05 | | 0.91 (0.88, 0.94) | <0.01 | | 0.88 (0.84, 0.92) | <0.01 |
| Coronary atherosclerosis and other heart disease | 0.90 (0.86, 0.94) | <0.01 | 1.28 (0.28, 5.90) | | >0.05 | 1.00 (0.88, 1.13) | >0.05 | | 0.94 (0.88, 0.99) | <0.05 | | 0.82 (0.77, 0.87) | <0.01 |
| Heart Failure and chronic rheumatic heart disease | 0.87 (0.81, 0.95) | <0.01 | 1.42 (0.23, 8.78) | | >0.05 | 1.02 (0.83, 1.26) | >0.05 | | 0.81 (0.71, 0.92) | <0.01 | | 0.87 (0.77, 0.98) | <0.05 |
| Hypertension | 0.89 (0.87, 0.91) | <0.01 | 1.95 (1.20, 3.16) | | <0.01 | 0.95 (0.90, 0.99) | <0.05 | | 0.87 (0.84, 0.90) | <0.01 | | 0.84 (0.80, 0.89) | <0.01 |
| Cardiovascular disease | 0.74 (0.55, 1.01) | >0.05 | 0.00 (0.00, 0.00) | | null | 0.76 (0.32, 1.81) | >0.05 | | 0.90 (0.59, 1.37) | >0.05 | | 0.55 (0.32, 0.93) | <0.05 |
| Cardiac dysrhythmia | 0.88 (0.85, 0.92) | <0.01 | 1.16 (0.70, 1.92) | | >0.05 | 0.89 (0.82, 0.97) | <0.05 | | 0.91 (0.86, 0.97) | <0.01 | | 0.81 (0.76, 0.87) | <0.01 |
| Acute myocardial infarction | 0.80 (0.73, 0.88) | <0.01 | 0.00 (0.00, 0.00) | | null | 0.90 (0.69, 1.18) | >0.05 | | 0.86 (0.75, 0.99) | <0.05 | | 0.71 (0.61, 0.82) | <0.01 |
| Myocarditis and cardiomyopathy | 0.82 (0.75, 0.89) | <0.01 | 2.25 (0.49, 10.31) | | >0.05 | 1.11 (0.90, 1.38) | >0.05 | | 0.82 (0.71, 0.93) | <0.01 | | 0.69 (0.60, 0.80) | <0.01 |
| **Respiratory** | **0.92 (0.91, 0.94)** | **<0.01** | **1.10 (0.98, 1.24)** | | **>0.05** | **0.98 (0.95, 1.01)** | **>0.05** | | **0.91 (0.88, 0.93)** | **<0.01** | | **0.85 (0.81, 0.88)** | **<0.01** |
| Respiratory symptoms and disease | 0.92 (0.90, 0.93) | <0.01 | 1.12 (1.00, 1.26) | | >0.05 | 0.96 (0.93, 0.99) | <0.05 | | 0.90 (0.88, 0.93) | <0.01 | | 0.85 (0.81, 0.88) | <0.01 |
| Acute pulmonary embolism | 0.75 (0.67, 0.85) | <0.01 | 2.19 (0.29, 16.29) | | >0.05 | 0.78 (0.60, 1.02) | >0.05 | | 0.77 (0.65, 0.91) | <0.01 | | 0.69 (0.56, 0.84) | <0.01 |
| Asthma | 1.05 (1.02, 1.09) | <0.01 | 1.34 (1.10, 1.62) | | <0.01 | 1.17 (1.11, 1.23) | <0.01 | | 0.99 (0.93, 1.04) | >0.05 | | 0.89 (0.81, 0.97) | <0.01 |
| **Renal** |  |  |  | |  |  |  | |  |  | |  |  |
| Kidney disease | 0.75 (0.72, 0.79) | <0.01 | 1.56 (0.55, 4.37) | | >0.05 | 0.86 (0.76, 0.97) | <0.05 | | 0.75 (0.70, 0.81) | <0.01 | | 0.69 (0.64, 0.73) | <0.01 |
| **Hemolytic and vascular** | **0.86 (0.82, 0.90)** | **<0.01** | **1.18 (0.53, 2.63)** | | **>0.05** | **0.99 (0.89, 1.10)** | **>0.05** | | **0.84 (0.79, 0.90)** | **<0.01** | | **0.81 (0.75, 0.86)** | **<0.01** |
| Coagulation and hemorrhagic disorders | 0.92 (0.87, 0.97) | <0.01 | 1.43 (0.61, 3.38) | | >0.05 | 1.05 (0.93, 1.19) | >0.05 | | 0.90 (0.83, 0.99) | <0.05 | | 0.84 (0.77, 0.91) | <0.01 |
| Thromboembolic event | 0.69 (0.62, 0.77) | <0.01 | 0.00 (0.00, Inf) | | >0.05 | 0.71 (0.56, 0.90) | <0.01 | | 0.65 (0.55, 0.77) | <0.01 | | 0.71 (0.60, 0.85) | <0.01 |
| Cerebrovascular disease | 0.83 (0.76, 0.89) | <0.01 | 0.36 (0.04, 3.49) | | >0.05 | 0.94 (0.72, 1.22) | >0.05 | | 0.83 (0.73, 0.94) | <0.01 | | 0.79 (0.71, 0.89) | <0.01 |
| **Gastrointestinal** | **0.94 (0.92, 0.96)** | **<0.01** | **0.90 (0.72, 1.12)** | | **>0.05** | **0.99 (0.95, 1.03)** | **>0.05** | | **0.94 (0.90, 0.97)** | **<0.01** | | **0.87 (0.83, 0.91)** | **<0.01** |
| Change in Bowel Habits | 0.91 (0.89, 0.94) | <0.01 | 0.89 (0.69, 1.15) | | >0.05 | 0.98 (0.93, 1.04) | >0.05 | | 0.90 (0.86, 0.94) | <0.01 | | 0.83 (0.78, 0.88) | <0.01 |
| Esophageal disorders | 0.95 (0.93, 0.98) | <0.01 | 0.87 (0.64, 1.18) | | >0.05 | 0.99 (0.95, 1.04) | >0.05 | | 0.96 (0.92, 0.99) | <0.05 | | 0.87 (0.83, 0.92) | <0.01 |
| **Neurologic** | **0.90 (0.88, 0.92)** | **<0.01** | **1.04 (0.90, 1.20)** | | **>0.05** | **0.96 (0.93, 1.00)** | **<0.05** | | **0.87 (0.85, 0.90)** | **<0.01** | | **0.85 (0.81, 0.88)** | **<0.01** |
| Ataxia / Trouble Walking | 0.83 (0.80, 0.87) | <0.01 | 0.97 (0.60, 1.57) | | >0.05 | 1.00 (0.91, 1.10) | >0.05 | | 0.83 (0.78, 0.88) | <0.01 | | 0.74 (0.69, 0.79) | <0.01 |
| Autonomic Dysfunction | 1.01 (0.90, 1.14) | >0.05 | 2.45 (0.59, 10.26) | | >0.05 | 1.07 (0.87, 1.32) | >0.05 | | 0.99 (0.82, 1.19) | >0.05 | | 0.93 (0.73, 1.19) | >0.05 |
| Cognitive disorders | 0.81 (0.72, 0.91) | <0.01 | 0.83 (0.28, 2.51) | | >0.05 | 0.90 (0.74, 1.10) | >0.05 | | 0.88 (0.74, 1.05) | >0.05 | | 0.61 (0.49, 0.77) | <0.01 |
| Headache | 0.93 (0.90, 0.95) | <0.01 | 0.98 (0.83, 1.15) | | >0.05 | 0.96 (0.92, 1.00) | <0.05 | | 0.90 (0.86, 0.94) | <0.01 | | 0.90 (0.84, 0.96) | <0.01 |
| Hearing loss and disturbances | 1.04 (0.98, 1.10) | >0.05 | 1.24 (0.75, 2.06) | | >0.05 | 1.25 (1.10, 1.41) | <0.01 | | 0.92 (0.84, 1.00) | >0.05 | | 1.04 (0.94, 1.15) | >0.05 |
| Myoneural Disorders | 0.99 (0.88, 1.12) | >0.05 | 1.58 (0.46, 5.47) | | >0.05 | 1.12 (0.89, 1.42) | >0.05 | | 1.03 (0.86, 1.22) | >0.05 | | 0.76 (0.58, 0.99) | <0.05 |
| Nervous System | 0.88 (0.78, 1.00) | <0.05 | 0.97 (0.31, 2.99) | | >0.05 | 0.97 (0.79, 1.18) | >0.05 | | 0.92 (0.76, 1.10) | >0.05 | | 0.64 (0.47, 0.87) | <0.01 |
| Peripheral Nerve Disorders | 0.95 (0.92, 0.99) | <0.01 | 0.95 (0.45, 1.99) | | >0.05 | 1.01 (0.94, 1.08) | >0.05 | | 0.93 (0.89, 0.98) | <0.01 | | 0.91 (0.84, 0.98) | <0.05 |
| Syncope and dizziness | 0.89 (0.86, 0.91) | <0.01 | 1.07 (0.85, 1.36) | | >0.05 | 0.89 (0.84, 0.94) | <0.01 | | 0.88 (0.84, 0.92) | <0.01 | | 0.86 (0.82, 0.92) | <0.01 |
| Visual disturbances | 1.00 (0.93, 1.07) | >0.05 | 1.52 (0.88, 2.63) | | >0.05 | 0.99 (0.88, 1.12) | >0.05 | | 0.94 (0.85, 1.05) | >0.05 | | 1.05 (0.91, 1.21) | >0.05 |
| Seizures | 0.73 (0.66, 0.82) | <0.01 | 0.93 (0.49, 1.79) | | >0.05 | 0.79 (0.67, 0.94) | <0.01 | | 0.72 (0.60, 0.88) | <0.01 | | 0.59 (0.47, 0.75) | <0.01 |
| Sensations and Perceptions | 0.77 (0.63, 0.95) | <0.05 | 0.00 (0.00, Inf) | | >0.05 | 0.68 (0.49, 0.94) | <0.05 | | 0.83 (0.59, 1.18) | >0.05 | | 0.87 (0.59, 1.30) | >0.05 |
| Smell and taste disturbances | 1.04 (0.85, 1.27) | >0.05 | 5.21 (0.47, 57.96) | | >0.05 | 1.05 (0.76, 1.45) | >0.05 | | 1.01 (0.74, 1.37) | >0.05 | | 0.99 (0.63, 1.56) | >0.05 |
| **Mental health** | **0.95 (0.93, 0.97)** | **<0.01** | **1.00 (0.85, 1.18)** | | **>0.05** | **0.99 (0.95, 1.02)** | **>0.05** | | **0.95 (0.92, 0.98)** | **<0.01** | | **0.89 (0.85, 0.93)** | **<0.01** |
| Sleeping conditions | 1.00 (0.97, 1.02) | >0.05 | 0.84 (0.63, 1.13) | | >0.05 | 1.03 (0.98, 1.07) | >0.05 | | 1.01 (0.97, 1.05) | >0.05 | | 0.91 (0.85, 0.96) | <0.01 |
| Other mental conditions | 0.79 (0.73, 0.86) | <0.01 | 0.84 (0.58, 1.22) | | >0.05 | 0.83 (0.73, 0.94) | <0.01 | | 0.79 (0.68, 0.91) | <0.01 | | 0.67 (0.54, 0.82) | <0.01 |
| Substance-related disorder | 0.68 (0.62, 0.74) | <0.01 | 0.50 (0.18, 1.38) | | >0.05 | 0.64 (0.57, 0.73) | <0.01 | | 0.70 (0.61, 0.80) | <0.01 | | 0.71 (0.58, 0.89) | <0.01 |
| Anxiety | 0.95 (0.93, 0.98) | <0.01 | 1.11 (0.94, 1.32) | | >0.05 | 1.00 (0.97, 1.04) | >0.05 | | 0.92 (0.88, 0.95) | <0.01 | | 0.89 (0.84, 0.95) | <0.01 |
| Mood disorder | 0.90 (0.87, 0.93) | <0.01 | 0.98 (0.79, 1.22) | | >0.05 | 0.92 (0.87, 0.96) | <0.01 | | 0.93 (0.88, 0.98) | <0.01 | | 0.81 (0.75, 0.88) | <0.01 |
| **Musculoskeletal** | **0.90 (0.89, 0.92)** | **<0.01** | **0.94 (0.82, 1.07)** | | **>0.05** | **0.95 (0.92, 0.98)** | **<0.01** | | **0.90 (0.87, 0.92)** | **<0.01** | | **0.85 (0.82, 0.88)** | **<0.01** |
| Malaise and fatigue | 0.93 (0.91, 0.95) | <0.01 | 1.06 (0.86, 1.30) | | >0.05 | 0.99 (0.95, 1.03) | >0.05 | | 0.93 (0.89, 0.96) | <0.01 | | 0.83 (0.79, 0.88) | <0.01 |
| Musculoskeletal pain | 0.92 (0.91, 0.94) | <0.01 | 0.93 (0.81, 1.07) | | >0.05 | 0.95 (0.92, 0.98) | <0.01 | | 0.91 (0.89, 0.94) | <0.01 | | 0.90 (0.87, 0.94) | <0.01 |
| Muscle disorder | 0.83 (0.80, 0.86) | <0.01 | 0.79 (0.50, 1.25) | | >0.05 | 0.95 (0.89, 1.02) | >0.05 | | 0.87 (0.82, 0.92) | <0.01 | | 0.65 (0.60, 0.70) | <0.01 |
| **Diabetes** | **0.90 (0.87, 0.93)** | **<0.01** | **1.43 (0.79, 2.61)** | | **>0.05** | **1.05 (0.97, 1.13)** | **>0.05** | | **0.89 (0.84, 0.94)** | **<0.01** | | **0.76 (0.70, 0.82)** | **<0.01** |
| Diabetes Type 1 | 0.91 (0.80, 1.03) | >0.05 | 0.94 (0.23, 3.79) | | >0.05 | 1.02 (0.80, 1.31) | >0.05 | | 0.93 (0.77, 1.12) | >0.05 | | 0.73 (0.56, 0.95) | <0.05 |
| Diabetes Type 2 | 0.91 (0.88, 0.95) | <0.01 | 1.74 (0.98, 3.09) | | >0.05 | 1.08 (1.00, 1.16) | <0.05 | | 0.90 (0.86, 0.95) | <0.01 | | 0.76 (0.70, 0.82) | <0.01 |
| **Other** | **0.85 (0.82, 0.87)** | **<0.01** | **0.91 (0.69, 1.21)** | | **>0.05** | **0.96 (0.90, 1.01)** | **>0.05** | | **0.87 (0.83, 0.92)** | **<0.01** | | **0.70 (0.66, 0.75)** | **<0.01** |
| Encephalitis | 0.24 (0.06, 1.03) | >0.05 | 0.00 (0.00, 0.00) | | null | 0.00 (0.00, Inf) | >0.05 | | 0.24 (0.03, 1.89) | >0.05 | | 0.32 (0.04, 2.59) | >0.05 |
| Weight Loss | 0.79 (0.75, 0.84) | <0.01 | 0.98 (0.66, 1.45) | | >0.05 | 0.86 (0.77, 0.97) | <0.05 | | 0.84 (0.77, 0.92) | <0.01 | | 0.67 (0.61, 0.73) | <0.01 |
| Rash | 1.05 (0.94, 1.17) | >0.05 | 0.38 (0.08, 1.72) | | >0.05 | 0.97 (0.81, 1.16) | >0.05 | | 1.13 (0.97, 1.32) | >0.05 | | 1.02 (0.74, 1.41) | >0.05 |
| PCC | 0.87 (0.82, 0.92) | <0.01 | 2.04 (0.97, 4.26) | | >0.05 | 1.07 (0.96, 1.19) | >0.05 | | 0.86 (0.78, 0.93) | <0.01 | | 0.68 (0.60, 0.77) | <0.01 |
| Symptoms and signs involving cognition, perception, emotional state and behavior | 0.81 (0.77, 0.85) | <0.01 | 0.74 (0.47, 1.17) | | >0.05 | 0.92 (0.84, 1.00) | >0.05 | | 0.84 (0.78, 0.91) | <0.01 | | 0.69 (0.63, 0.74) | <0.01 |

Abbreviations: PCC – Post-COVID Conditions

^a^Hazard ratios were adjusted for age in years (continuous), sex, HHS region, obesity, number of underlying risk factors (1, 2, 3, 4, or ≥5), SARS-CoV-2 vaccination history (any vaccination ≥14 days prior to index date vs. no vaccination), and number of healthcare encounters in the year prior to index date

Supplemental Table 7. Adjusted^a^ hazard ratios for Post-COVID Conditions (PCC) excluding patients with a nirmatrelvir-ritonavir COVID-19 index event, by age group, among persons treated with nirmatrelvir-ritonavir during acute COVID-19 illness compared to untreated, Health Verity, April 1-December 31, 2022.

|  | **Overall (all ages)** | | **Ages 12-17** | | **Ages 18-49** | | **Ages 50-64** | | **Ages 65+** | |
| --- | --- | --- | --- | --- | --- | --- | --- | --- | --- | --- |
|  | **Hazard Ratio (95% CI)** | **p** | **Hazard Ratio (95% CI)** | **p** | **Hazard Ratio (95% CI)** | **p** | **Hazard Ratio (95% CI)** | **p** | **Hazard Ratio (95% CI)** | **p** |
| **Experienced post-COVID-19 symptoms/conditions** |  |  |  |  |  |  |  |  |  |  |
| At least 1 symptom/condition | 1.01 (1.00, 1.02) | <0.01 | 1.08 (1.00, 1.16) | <0.05 | 1.05 (1.04, 1.07) | <0.01 | 1.02 (1.01, 1.04) | <0.01 | 0.92 (0.90, 0.94) | <0.01 |
| ≥2 symptoms/conditions | 0.97 (0.96, 0.99) | <0.01 | 1.08 (0.95, 1.23) | >0.05 | 1.04 (1.01, 1.06) | <0.01 | 0.99 (0.97, 1.01) | >0.05 | 0.84 (0.82, 0.87) | <0.01 |
| **Post-COVID Conditions** |  |  |  |  |  |  |  |  |  |  |
| **Cardiovascular** | **0.99 (0.97, 1.02)** | **>0.05** | **1.21 (0.96, 1.52)** | **>0.05** | **1.04 (0.99, 1.09)** | **>0.05** | **1.00 (0.96, 1.04)** | **>0.05** | **0.85 (0.79, 0.90)** | **<0.01** |
| Circulatory signs and symptoms | 1.00 (0.97, 1.03) | >0.05 | 1.32 (1.03, 1.69) | <0.05 | 1.04 (0.99, 1.09) | >0.05 | 1.00 (0.95, 1.04) | >0.05 | 0.90 (0.84, 0.96) | <0.01 |
| Coronary atherosclerosis and other heart disease | 0.90 (0.86, 0.95) | <0.01 | -- |  | 0.95 (0.82, 1.10) | >0.05 | 0.99 (0.92, 1.06) | >0.05 | 0.78 (0.72, 0.85) | <0.01 |
| Heart Failure and chronic rheumatic heart disease | 0.88 (0.80, 0.98) | <0.05 | 2.83 (0.18, 45.21) | >0.05 | 0.93 (0.73, 1.20) | >0.05 | 0.85 (0.72, 1.00) | <0.05 | 0.89 (0.76, 1.04) | >0.05 |
| Hypertension | 0.94 (0.91, 0.97) | <0.01 | 1.60 (0.94, 2.74) | >0.05 | 0.98 (0.92, 1.04) | >0.05 | 0.94 (0.90, 0.99) | <0.05 | 0.85 (0.79, 0.90) | <0.01 |
| Cardiovascular disease | 0.65 (0.42, 0.99) | <0.05 | -- |  | 0.44 (0.13, 1.53) | >0.05 | 0.80 (0.45, 1.43) | >0.05 | 0.53 (0.26, 1.10) | >0.05 |
| Cardiac dysrhythmia | 0.89 (0.85, 0.94) | <0.01 | 1.18 (0.62, 2.21) | >0.05 | 0.95 (0.86, 1.07) | >0.05 | 0.91 (0.84, 0.99) | <0.05 | 0.79 (0.72, 0.86) | <0.01 |
| Acute myocardial infarction | 0.77 (0.68, 0.87) | <0.01 | -- |  | 0.82 (0.59, 1.14) | >0.05 | 0.88 (0.73, 1.06) | >0.05 | 0.65 (0.53, 0.79) | <0.01 |
| Myocarditis and cardiomyopathy | 0.86 (0.77, 0.96) | <0.01 | 0.70 (0.13, 3.85) | >0.05 | 1.21 (0.93, 1.57) | >0.05 | 0.86 (0.72, 1.01) | >0.05 | 0.72 (0.61, 0.86) | <0.01 |
| **Respiratory** | 1.07 (1.05, 1.10) | **<0.01** | **1.14 (0.99, 1.31)** | **>0.05** | **1.14 (1.10, 1.18)** | **<0.01** | **1.08 (1.04, 1.11)** | **<0.01** | **0.94 (0.89, 0.99)** | **<0.05** |
| Respiratory symptoms and disease | 1.06 (1.04, 1.08) | <0.01 | 1.17 (1.02, 1.34) | <0.05 | 1.11 (1.07, 1.15) | <0.01 | 1.07 (1.03, 1.10) | <0.01 | 0.95 (0.90, 1.00) | <0.05 |
| Acute pulmonary embolism | 0.75 (0.65, 0.86) | <0.01 | -- |  | 0.77 (0.57, 1.05) | >0.05 | 0.73 (0.59, 0.90) | <0.01 | 0.69 (0.53, 0.90) | <0.01 |
| Asthma | 1.17 (1.12, 1.22) | <0.01 | 1.31 (1.05, 1.64) | <0.05 | 1.34 (1.26, 1.43) | <0.01 | 1.10 (1.03, 1.18) | <0.01 | 0.88 (0.79, 0.99) | <0.05 |
| **Renal** |  |  |  |  |  |  |  |  |  |  |
| Kidney disease | 0.77 (0.73, 0.82) | <0.01 | 0.98 (0.28, 3.44) | >0.05 | 0.85 (0.73, 0.99) | <0.05 | 0.78 (0.71, 0.86) | <0.01 | 0.70 (0.64, 0.76) | <0.01 |
| **Hemolytic and vascular** | **0.88 (0.84, 0.94)** | **<0.01** | **0.89 (0.33, 2.41)** | **>0.05** | **1.06 (0.94, 1.20)** | **>0.05** | **0.84 (0.77, 0.92)** | **<0.01** | **0.83 (0.76, 0.91)** | **<0.01** |
| Coagulation and hemorrhagic disorders | 0.92 (0.86, 0.99) | <0.05 | 1.01 (0.37, 2.78) | >0.05 | 1.11 (0.96, 1.29) | >0.05 | 0.87 (0.78, 0.97) | <0.05 | 0.86 (0.77, 0.97) | <0.05 |
| Thromboembolic event | 0.73 (0.64, 0.84) | <0.01 | -- |  | 0.79 (0.59, 1.04) | >0.05 | 0.71 (0.58, 0.88) | <0.01 | 0.70 (0.56, 0.88) | <0.01 |
| Cerebrovascular disease | 0.85 (0.76, 0.94) | <0.01 | -- |  | 1.01 (0.73, 1.39) | >0.05 | 0.79 (0.67, 0.94) | <0.01 | 0.84 (0.73, 0.98) | <0.05 |
| **Gastrointestinal** | **1.00 (0.98, 1.03)** | **>0.05** | **0.90 (0.68, 1.19)** | **>0.05** | **1.07 (1.02, 1.12)** | **<0.01** | **0.99 (0.95, 1.04)** | **>0.05** | **0.92 (0.87, 0.98)** | **<0.05** |
| Change in Bowel Habits | 0.99 (0.95, 1.03) | >0.05 | 1.03 (0.75, 1.41) | >0.05 | 1.06 (1.00, 1.13) | >0.05 | 0.97 (0.91, 1.03) | >0.05 | 0.90 (0.83, 0.98) | <0.05 |
| Esophageal disorders | 1.02 (0.99, 1.05) | >0.05 | 0.88 (0.59, 1.32) | >0.05 | 1.09 (1.03, 1.15) | <0.01 | 1.02 (0.97, 1.06) | >0.05 | 0.91 (0.85, 0.98) | <0.01 |
| **Neurologic** | **0.98 (0.96, 1.01)** | **>0.05** | **1.07 (0.90, 1.26)** | **>0.05** | **1.06 (1.02, 1.11)** | **<0.01** | **0.97 (0.93, 1.00)** | **>0.05** | **0.88 (0.83, 0.93)** | **<0.01** |
| Ataxia / Trouble Walking | 0.89 (0.84, 0.93) | <0.01 | 1.31 (0.67, 2.57) | >0.05 | 1.04 (0.93, 1.16) | >0.05 | 0.92 (0.85, 1.00) | <0.05 | 0.77 (0.70, 0.83) | <0.01 |
| Autonomic Dysfunction | 0.98 (0.84, 1.13) | >0.05 | 1.54 (0.31, 7.66) | >0.05 | 1.01 (0.79, 1.29) | >0.05 | 0.91 (0.72, 1.16) | >0.05 | 1.03 (0.76, 1.39) | >0.05 |
| Cognitive disorders | 0.83 (0.72, 0.95) | <0.01 | 0.82 (0.21, 3.22) | >0.05 | 0.90 (0.72, 1.13) | >0.05 | 0.93 (0.75, 1.15) | >0.05 | 0.60 (0.45, 0.80) | <0.01 |
| Headache | 1.03 (1.00, 1.06) | >0.05 | 1.01 (0.83, 1.22) | >0.05 | 1.06 (1.01, 1.12) | <0.05 | 1.01 (0.95, 1.06) | >0.05 | 0.99 (0.90, 1.08) | >0.05 |
| Hearing loss and disturbances | 1.07 (1.00, 1.15) | >0.05 | 1.04 (0.57, 1.90) | >0.05 | 1.28 (1.10, 1.48) | <0.01 | 0.97 (0.87, 1.08) | >0.05 | 1.06 (0.94, 1.21) | >0.05 |
| Myoneural Disorders | 0.94 (0.81, 1.10) | >0.05 | 1.47 (0.24, 8.78) | >0.05 | 1.12 (0.85, 1.47) | >0.05 | 0.97 (0.78, 1.21) | >0.05 | 0.67 (0.47, 0.95) | <0.05 |
| Nervous System | 0.89 (0.77, 1.03) | >0.05 | 0.82 (0.21, 3.22) | >0.05 | 0.96 (0.76, 1.21) | >0.05 | 0.97 (0.77, 1.22) | >0.05 | 0.56 (0.37, 0.85) | <0.01 |
| Peripheral Nerve Disorders | 0.99 (0.95, 1.04) | >0.05 | 0.64 (0.23, 1.80) | >0.05 | 1.04 (0.96, 1.13) | >0.05 | 1.00 (0.94, 1.07) | >0.05 | 0.90 (0.82, 1.00) | <0.05 |
| Syncope and dizziness | 0.97 (0.94, 1.01) | >0.05 | 1.20 (0.90, 1.59) | >0.05 | 1.01 (0.95, 1.08) | >0.05 | 0.97 (0.92, 1.03) | >0.05 | 0.89 (0.82, 0.96) | <0.01 |
| Visual disturbances | 1.12 (1.03, 1.22) | <0.05 | 1.29 (0.67, 2.50) | >0.05 | 1.09 (0.94, 1.26) | >0.05 | 1.11 (0.97, 1.27) | >0.05 | 1.14 (0.95, 1.36) | >0.05 |
| Seizures | 0.73 (0.64, 0.84) | <0.01 | 0.76 (0.31, 1.88) | >0.05 | 0.85 (0.69, 1.05) | >0.05 | 0.68 (0.54, 0.86) | <0.01 | 0.58 (0.43, 0.77) | <0.01 |
| Sensations and Perceptions | 0.85 (0.66, 1.09) | >0.05 | -- |  | 0.87 (0.58, 1.29) | >0.05 | 0.75 (0.48, 1.16) | >0.05 | 1.00 (0.62, 1.64) | >0.05 |
| Smell and taste disturbances | 1.09 (0.84, 1.40) | >0.05 | -- |  | 0.94 (0.63, 1.41) | >0.05 | 1.10 (0.75, 1.62) | >0.05 | 1.34 (0.70, 2.56) | >0.05 |
| **Mental health** | **1.05 (1.03, 1.08)** | **<0.01** | **1.16 (0.95, 1.41)** | **>0.05** | **1.08 (1.04, 1.13)** | **<0.01** | **1.08 (1.04, 1.13)** | **<0.01** | **0.92 (0.86, 0.98)** | **<0.01** |
| Sleeping conditions | 1.06 (1.03, 1.10) | <0.01 | 0.91 (0.64, 1.31) | >0.05 | 1.08 (1.02, 1.14) | <0.05 | 1.09 (1.04, 1.15) | <0.01 | 0.95 (0.88, 1.02) | >0.05 |
| Other mental conditions | 0.85 (0.77, 0.94) | <0.01 | 0.96 (0.63, 1.47) | >0.05 | 0.89 (0.77, 1.03) | >0.05 | 0.89 (0.74, 1.05) | >0.05 | 0.64 (0.49, 0.84) | <0.01 |
| Substance-related disorder | 0.72 (0.65, 0.80) | <0.01 | 0.67 (0.24, 1.86) | >0.05 | 0.68 (0.58, 0.79) | <0.01 | 0.78 (0.67, 0.91) | <0.01 | 0.70 (0.53, 0.92) | <0.05 |
| Anxiety | 1.05 (1.02, 1.08) | <0.01 | 1.40 (1.13, 1.73) | <0.01 | 1.10 (1.05, 1.15) | <0.01 | 1.04 (0.99, 1.09) | >0.05 | 0.90 (0.83, 0.97) | <0.05 |
| Mood disorder | 0.94 (0.90, 0.98) | <0.01 | 0.97 (0.75, 1.25) | >0.05 | 0.93 (0.87, 0.99) | <0.05 | 0.99 (0.92, 1.06) | >0.05 | 0.85 (0.77, 0.95) | <0.01 |
| **Musculoskeletal** | **1.01 (0.99, 1.03)** | **>0.05** | **0.92 (0.78, 1.08)** | **>0.05** | **1.04 (1.01, 1.08)** | **<0.05** | **1.03 (1.00, 1.07)** | **<0.05** | **0.90 (0.86, 0.95)** | **<0.01** |
| Malaise and fatigue | 1.01 (0.98, 1.04) | >0.05 | 1.07 (0.83, 1.37) | >0.05 | 1.08 (1.03, 1.13) | <0.01 | 1.02 (0.97, 1.07) | >0.05 | 0.87 (0.82, 0.93) | <0.01 |
| Musculoskeletal pain | 1.02 (1.00, 1.05) | <0.05 | 0.92 (0.77, 1.10) | >0.05 | 1.04 (1.00, 1.08) | <0.05 | 1.05 (1.01, 1.08) | <0.01 | 0.95 (0.90, 1.00) | <0.05 |
| Muscle disorder | 0.87 (0.83, 0.91) | <0.01 | 0.73 (0.40, 1.33) | >0.05 | 1.01 (0.93, 1.10) | >0.05 | 0.91 (0.85, 0.97) | <0.01 | 0.65 (0.59, 0.71) | <0.01 |
| **Diabetes** | **0.93 (0.89, 0.97)** | **<0.01** | **1.28 (0.63, 2.62)** | **>0.05** | **1.04 (0.96, 1.14)** | **>0.05** | **0.94 (0.88, 1.01)** | **>0.05** | **0.76 (0.69, 0.84)** | **<0.01** |
| Diabetes Type 1 | 0.85 (0.72, 0.99) | <0.05 | 0.61 (0.12, 3.09) | >0.05 | 0.98 (0.72, 1.33) | >0.05 | 0.86 (0.68, 1.08) | >0.05 | 0.68 (0.48, 0.96) | <0.05 |
| Diabetes Type 2 | 0.94 (0.90, 0.99) | <0.05 | 1.92 (0.94, 3.93) | >0.05 | 1.08 (0.99, 1.17) | >0.05 | 0.95 (0.89, 1.02) | >0.05 | 0.75 (0.68, 0.83) | <0.01 |
| **Other** | **0.91 (0.87, 0.94)** | **<0.01** | **0.95 (0.68, 1.33)** | **>0.05** | **1.02 (0.95, 1.09)** | **>0.05** | **0.94 (0.89, 1.00)** | **<0.05** | **0.75 (0.70, 0.81)** | **<0.01** |
| Encephalitis | 0.27 (0.03, 2.19) | >0.05 | -- |  | -- |  | 0.46 (0.05, 3.99) | >0.05 | -- |  |
| Weight Loss | 0.77 (0.72, 0.83) | <0.01 | 0.84 (0.52, 1.35) | >0.05 | 0.85 (0.73, 0.97) | <0.05 | 0.80 (0.71, 0.89) | <0.01 | 0.66 (0.58, 0.75) | <0.01 |
| Rash | 1.19 (1.04, 1.36) | <0.05 | -- |  | 1.24 (1.00, 1.53) | >0.05 | 1.23 (1.01, 1.49) | <0.05 | 0.98 (0.64, 1.49) | >0.05 |
| PCC | 1.04 (0.97, 1.12) | >0.05 | 2.18 (1.00, 4.75) | >0.05 | 1.21 (1.07, 1.37) | <0.01 | 1.04 (0.93, 1.15) | >0.05 | 0.87 (0.75, 1.01) | >0.05 |
| Symptoms and signs involving cognition, perception, emotional state and behavior | 0.86 (0.81, 0.91) | <0.01 | 1.00 (0.58, 1.71) | >0.05 | 0.97 (0.87, 1.07) | >0.05 | 0.87 (0.79, 0.96) | <0.01 | 0.73 (0.66, 0.81) | <0.01 |

Abbreviations: PCC – Post-COVID Conditions

^a^Hazard ratios were adjusted for age in years (continuous), sex, HHS region, obesity, number of underlying risk factors (1, 2, 3, 4, or ≥5), SARS-CoV-2 vaccination history (any vaccination ≥14 days prior to index date vs. no vaccination), and number of healthcare encounters in the year prior to index date
